# MyGESig: a population-specific gene signature improves survival prediction in Malaysian breast cancer patients

**DOI:** 10.1101/2025.08.28.25334111

**Authors:** Mohamad Hazwan Khairi, Zhi Lei Wong, Boon Hong Ang, James Phipps-Tan, Putri Nur Fatin, Rajadurai Pathmanathan, See Mee Hoong, Nur Aishah Mohd Taib, Cheng-Har Yip, Weang Kee Ho, Mei Chee Tai, Soo-Hwang Teo, Sok Ching Cheong, Pan Jia-Wern

## Abstract

Accurate prognostic models are essential for guiding treatment decisions and improving patient outcomes in breast cancer. To achieve this, population-specific models are needed to account for genetic, clinical, and pathological differences across populations. In this study, the widely used and freely available PREDICT v3.0 breast cancer prognostic model was first validated in the multiethnic Malaysian Breast Cancer (MyBrCa) cohort to assess its performance. Given its only moderate performance in this population, a machine learning workflow was developed to integrate gene expression and clinical information for classifying patients by their 10-year prognosis. A 77-gene signature, termed MyGESig, was derived from the transcriptomes of 258 MyBrCa patients. Using this signature in combination with clinical variables, an ensemble-based model achieved a median area under the receiver-operator characteristic curve (AUROC) of 0.92 in the hold-out testing set and 0.90 in the independent MyBrCa dataset. While the model exhibited poor generalizability in external cohorts, its discriminative performance improved when trained and tested within the same population (median AUROC: 0.71 in METABRIC; 0.84 in SCAN-B), validating the prognostic value of the gene set. Together, these findings demonstrate the value of incorporating population-specific gene expression datasets into prognosis prediction and highlight the need to develop and validate models tailored to diverse populations in breast cancer.

## 2 Introduction

Prognostic tools are essential in the management of breast cancer as they provide personalized risk assessments to guide treatment decisions, optimize adjuvant therapy selection, and improve survival outcomes while avoiding overtreatment in low-risk patients (Cardoso et al., 2019). One such tool is PREDICT: a widely used, freely available web-based prognostic tool that estimates survival outcomes and treatment benefits (e.g., chemotherapy, endocrine therapy, anti-HER2 therapy) in breast cancer patients by integrating clinicopathological factors such as age, tumor size, grade, and receptor status (Wishart et al., 2010). PREDICT version 3.0 has been released, with the model being refitted on an updated version of its original cohort, now including a larger number of patients, an improved prognosis data for early breast cancer, and a higher proportion of ER-negative cases (Grootes et al., 2024). PREDICT v3.0 has significantly influenced clinical decision-making, particularly in adjuvant therapy planning, and is highly accurate in women of European descent in North America and Europe (Gray et al., 2018; Stabellini et al., 2023). Other prognostic tools include genomic signatures such as Oncotype DX, MammaPrint, and Prosigna, which are molecular assays that analyse tumor gene expression patterns to predict recurrence risk and chemotherapy benefit in breast cancer, enabling personalized treatment decisions that reduce overtreatment in low-risk patients while identifying high-risk cases needing aggressive therapy (Paik et al., 2004; Vijver et al., 2002; Wallden et al., 2015).

In women of Asian descent, the accuracy of the PREDICT tool and genomic signatures is lower than that in women of European descent, presumably because of population-specific differences in tumor biology and treatment response. Asian breast cancer patients, for instance, are younger at diagnosis and have a higher proportion of premenopausal, oestrogen receptor (ER) negative and human epidermal growth factor receptor 2 (HER2) receptor-positive cases compared to their Western counterparts (Yap et al., 2019). Furthermore, previous findings revealed that the molecular landscape of Asian breast cancer patients is significantly different from that of Caucasian women, characterized by elevated expression of immune-related gene signatures and higher prevalence of TNBC and HER2-enriched molecular subtype (Lee et al., 2024; Pan et al., 2020). Indeed, the earlier versions of PREDICT have been evaluated in Malaysia, India, and Japan, with the reported AUROC for 5-year estimation of overall survival being 0.78, 0.64, 0.71, and compared to 0.78 in women in the USA and 0.80 in the Netherlands (Nair et al., 2023; Stabellini et al., 2023; van Maaren et al., 2017; Wong et al., 2015; Zaguirre et al., 2021). However, despite good discrimination overall, poor calibration was observed in certain subgroups across different cohorts, including younger patients (< 40 years old), older patients (≥ 65 years old), and those receiving specific treatment regimens, which likely reflect the differences in patient characteristics compared to the original cohort. The Western-centric development of these models further compounds this issue and contributes to their poorer performance in Asian cohorts. Similarly, when the 21-gene recurrence score (Oncotype DX) predicted recurrence risk in HR+/HER2- patients in Japan and South Korea, a higher proportion of patients were classified as intermediate risk compared to cohorts in North America and Europe (Kim et al., 2018; Toi et al., 2010). Given these population-specific differences, several multi-gene assays have been developed specifically for women of East Asian descent. For instance, the GenesWell BCT assay, developed from Korean patient data, was able to classify more patients under 50 years old as low-risk than the Oncotype DX test (Kwon et al., 2019). In Taiwanese women, a 34-gene panel was developed to classify patients into low- and high-risk groups for local/regional recurrence (LRR) after mastectomy (Cheng et al., 2016).

Earlier validations of PREDICT in Malaysian populations were conducted on smaller cohorts and relied on an older version of the model (v2.2). In this study, the performance of PREDICT v3.0 was assessed by validating it in the MyBrCa cohort, a large cohort of Malaysian breast cancer patients, including women of Chinese, Malay, and Indian ethnicities. To address the need for a population-specific prognostic model, a machine learning framework was developed that incorporates gene expression and clinical data to classify patients by 10-year prognosis. Through this framework, a 77-gene signature, termed MyGESig, was derived from the transcriptome of 258 MyBrCa patients. The predictive ability of this signature was then evaluated by comparing its discriminative performance across different datasets and assessing its prognostic value.

## 3 Method

### 3.1 Study cohort

The clinical and gene expression data used in this study were derived from the MyBrCa, a hospital-based case-control study that was started in 2002 in two hospitals, which now includes a total of 6086 cases diagnosed sequentially from October 2002 to December 2016 (Tan et al., 2018). For the validation of PREDICT v3.0, a cohort was defined consisting of patients with complete clinical information required by the PREDICT v3.0 model, specifically tumor grade, age at diagnosis, tumor size, number of positive lymph nodes, and ER status (Figure 1).

**Figure 1.**
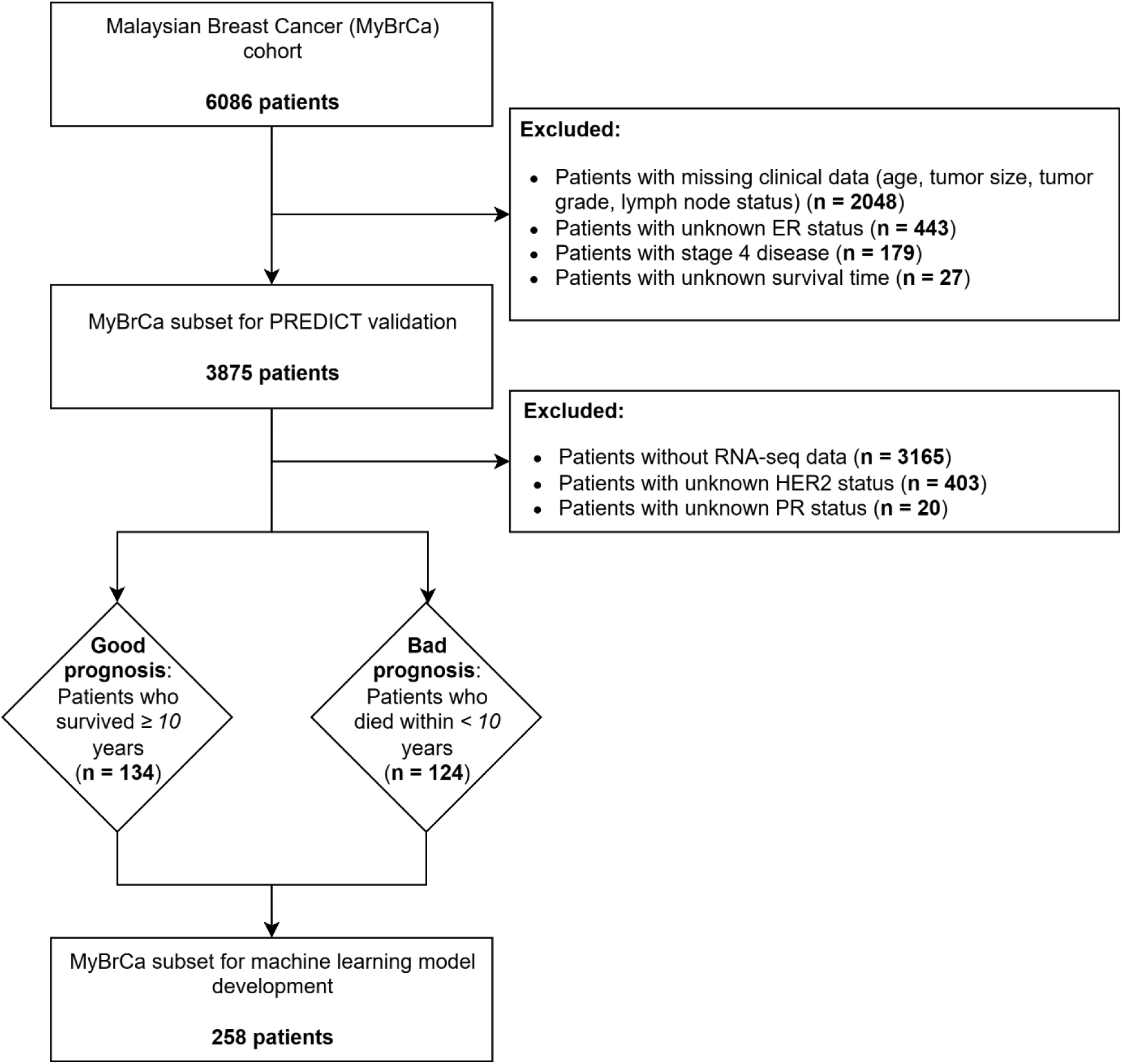
Flowchart summarizing cohort selection for PREDICT validation (n = 3875) and machine learning model development (n = 258)

HER2 and PR status were not required for inclusion, as the PREDICT model can accommodate unknown values for these markers. Patients with incomplete survival data, including vital status and survival time, were excluded. Survival time is calculated as the difference between the date of diagnosis and the date of last follow-up. Patients with stage 4 disease were also excluded from this cohort.

A separate cohort was defined for the development of a machine learning-based prognostic model. This cohort was a subset of the former, selected based on the availability of RNA-seq data. Unlike the PREDICT model, the machine learning model required complete HER2 and PR status, and therefore, patients missing either of these variables were excluded. Since the prognosis prediction task were addressed as a two-class classification task, patients were classified into good or poor prognosis groups using a 10-year survival threshold, as adapted from previous studies (Bertucci et al., 2002; Chen et al., 2012; Zhu et al., 2009). Specifically, those who survived more than 10 years after diagnosis were considered to have a good prognosis, whereas those who died within 10 years were classified as having a poor prognosis. Accordingly, patients who are still alive but had less than 10 years of follow-up were excluded to ensure accurate classification of prognosis.

### 3.2 Validation of PREDICT v3.0 in the MyBrCa cohort

Survival estimates at 5, 10, and 15 years were computed using the PREDICT v3.0 tool [GitHub (https://github.com/pengpclab/PREDICTv3)]. This version of PREDICT v3.0 requires input variables including age at diagnosis, tumor size, number of positive lymph nodes, ER status, tumor grade, mode of detection, and Ki67 status. HER2 status, PR status, smoking status, and treatment-related variables such as chemotherapy, hormone therapy, trastuzumab, and bisphosphonates are optional inputs and can be entered as unknown (coded as 9) when missing. Accounting for any treatments received, the predicted survival was obtained by summing the survival probabilities (post-surgery) and the calculated survival benefit from the treatments, as output by the PREDICT v3.0 model. The performance of PREDICT v3.0 was evaluated based on goodness-of-fit, calibration and discrimination. Goodness-of-fit was assessed graphically by plotting overall survival (calculated via Kaplan-Meier analysis) against the median predicted survival, with predictions grouped into quintiles. Model calibration was assessed by comparing the observed 5-, 10-, and 15-year overall survival (calculated via Kaplan-Meier analysis) versus the median probability of survival estimated by PREDICT v3.0. Model discrimination was assessed by calculating the AUROC curve using the pROC (v1.18.5) package (Robin et al., 2011). Subgroup analysis was performed according to age, ethnicity, stage, tumor grade, clinical subtype, receptor status, and systemic therapy received.

### 3.3 Gene expression dataset

Of the 6086 cases diagnosed sequentially from October 2002 to December 2016, 710 cases had fresh frozen tumor samples surplus to diagnostic requirements collected at surgery (Pan et al., 2021; Pan et al., 2020). Only 258 samples were included in this study, based on the availability of complete clinical information and eligibility under the prognosis classification criteria. mRNA extracted from breast tissue samples was subjected to paired-end 75 base pair RNA-Seq on the HiSeq4000 platform (Illumina, San Diego, USA). Details of sample collection, sequencing, and post-sequencing data processing were previously described in Pan et al. (2020). Clean count data were obtained by filtering out genes with low expression levels (in counts per million, CPM) using the filterByExpr() function from the edgeR R package (v4.0.16) (Chen et al., 2025). A gene was retained if its CPM exceeded a library-size-adjusted threshold in enough samples from at least one group. After filtering, 18,112 genes were retained and were then normalized using the trimmed mean of M-values (TMM) method from the edgeR (v4.0.16) R package. The normalized count data were subjected to voom transformation using the Limma (v3.58.1) R package (Ritchie et al., 2015) to prepare for downstream linear modelling analysis.

### 3.4 Identification of multigene prognostic signature

To reduce the complexity of the expression data and extract the most informative genes, principal component analysis (PCA) was applied to all samples. Principal components (PCs) that together accounted for 80% of the total variance were retained, and the top 30 genes with the highest loading scores from each of these PCs were extracted. This approach assumes that genes whose expression correlates with patient outcome should not be equal in expression across all patients in the dataset. If a gene has nearly the same expression level (or follows the same eigenvector of expression) across all patients, it is unlikely to carry meaningful information about differences in prognosis. By using PCA, we sought to eliminate such genes and retain only genes that contribute the most to variation in expression across the dataset.

To select relevant genes that are associated with prognosis, LASSO regression from the glmnet (v4.1-8) R package (Tay et al., 2023) was implemented. LASSO regression applies a penalty term proportional to the absolute value of the coefficients, shrinking their magnitude and thereby reducing the number of features in a model. This improves model performance by shrinking the coefficients of irrelevant features towards zero. To determine the optimal penalization strength, the Akaike Information Criterion (AIC) and the Bayesian Information Criterion (BIC) were used. AIC is a model selection criterion that evaluates the goodness of fit of different models and selects the best one by balancing model fit and complexity. The best-fit model, according to AIC, should explain the data well using the fewest number of independent variables. Similarly, BIC also considers model fit and complexity, but implements a stronger penalty for model complexity, thus favouring simpler models.

### 3.5 Gene set enrichment analysis

Gene ontology (GO) term annotation was performed using the clusterProfiler (v4.10.1) R package (Wu et al., 2021) to identify the biological processes (BP) and molecular functions (MF) terms associated with the gene signature. The overrepresentation of specific GO categories was assessed through gene set enrichment analysis using the gseGO() function. Additionally, pathway-level enrichment was also investigated using the GSEA() function and the hallmark gene set (H) (Liberzon et al., 2015) from the Molecular Signatures Database (MSigDB) (Subramanian et al., 2005).

### 3.6 Prediction of disease prognosis using machine learning

The prognostic model was developed in R (v4.3.3) using the machine learning framework available in the caret (v7.0-1) package (Kuhn, 2008). In brief, the framework consists of an outer loop for dataset splitting and an inner loop that combines pipelines based on Random Forest (RF) and Support Vector Machine (SVM) algorithms to predict the probability of a patient having a good prognosis (Supplementary Figure 1). Three SVM algorithms were used in this framework: linear, radial, and polynomial kernels. The predictions from them were averaged to get a consensus. The outer loop performs five iterations of different random seeds, where each iteration applies a one-fold random stratified split of the data in a 7:3 ratio, resulting in five distinct training and testing datasets. Within the inner loop, each of the distinct training datasets undergoes z-score normalization and collinearity removal before model training.

Default tuning parameters for each model were adopted during hyperparameter tuning, with a five-fold cross-validation (CV) employed to search for parameters that maximize the area under the receiver operating characteristic (AUROC) plot. The overall model is composed of five sets of five RF and SVM sub-models, resulting in a total of 25 sub-models for each algorithm. Within each set, the classification probabilities from all algorithms were averaged to achieve a robust prediction. The performance of the model was assessed by calculating the AUROC, and the model with the median AUROC across all hold-out testing datasets was selected as the final model. Other performance metrics, such as the F1 score, precision, recall, and Cohen’s kappa, were calculated using the classification threshold that maximized the Youden’s Index, as determined in the pROC (v1.18.5) package. Youden’s Index was chosen as it balances specificity and sensitivity, making it suitable for this prognostic model where accurate distinction between good and poor prognosis is equally important.

### 3.7 Validation on independent cohorts

In brief, the performance of the model was assessed in hold-out testing datasets and validation datasets from three independent cohorts: a subset of the MyBrCa cohort that was excluded from the initial stratified training-testing split, and two external cohorts (the METABRIC and the SCAN-B cohorts). The independent MyBrCa dataset was established by applying the same 7:3 stratified split used to generate training and testing datasets: the first 30% was retained as an independent validation dataset, while the remaining 70% was used to generate the training and testing datasets (Supplementary Figure 1). The clinical and gene expression data for both external cohorts were obtained from the cBioPortal (Cerami et al., 2012), and the same criteria were applied to exclude patients and classify them based on their 10-year survival. The gene expression datasets of both cohorts were cleaned and normalized as described in their respective publications (Curtis et al., 2012), and the SCAN-B RNA-Seq dataset was voom-transformed to facilitate linear modelling.

### 3.8 Prognostic evaluation using Cox proportional hazard model and survival analysis

Univariable Cox proportional hazard model (using the survival (v3.8-3) package (Borgan, 2001)) was used to assess the prognostic value of the machine learning model. This evaluation was performed on the independent MyBrCa dataset to prevent information leakage from the model training step. Predicted prognosis (classified as good or poor) from the machine learning model with median AUROC was used as an independent predictor in the Cox model. For comparison, the prognostic value of PREDICT v3.0 and established clinical factors, including stage, grade, hormone receptor positivity, and HER2 positivity, were also assessed using Cox model. The PREDICT v3.0 overall survival estimates were dichotomized using the classification threshold that maximized the Youden’s index, as determined by the pROC (v1.18.5) package. To assess whether the gene signature provides prognostic information independent of the clinical information, predicted prognosis from a model trained solely on gene expression data was included as a covariate in a multivariable model alongside the clinical factors that were found to be significant in the univariable analyses. The hazard ratios (HR) associated with the univariable models were plotted in a single forest plot using the ggplot2 (v3.5.1) package. Additionally, Kaplan–Meier survival curves were also constructed to compare the survival differences between prognosis groups predicted by the model and PREDICT v3.0.

## 4 Results

### 4.1 Cohort description

MyBrCa cohort was previously described in Tan et. al. (2018). For validation of PREDICT v 3.0, 3875 patients (out of 6086 patients) with complete clinical information required by the model were included. The median age at diagnosis of this subset of women was 51 years old (inter-quartile range, IQR: 45-60 years) (Table 1). The patients were followed for a median of 7.3 years, during which 709 deaths occurred, accounting for 18.3% of the cohort over the entire follow-up period. This cohort included a high proportion of ER-positive (69.9%) and PR-positive (61.7%) cases, and most of the patients were diagnosed with early-stage disease (Stage 0-2) (81.3%).

**Table 1.** Baseline clinicopathological characteristics of the MyBrCa, METABRIC, and SCAN-B cohorts used in this study. Reported variables include patient demographics, tumor biology (ER, PR, HER2 status), tumor stage and grade, tumor size, nodal involvement, and treatment details. MyBrCa = Malaysian Breast Cancer; METABRIC = Molecular Taxonomy of Breast Cancer International Consortium; SCAN-B = Sweden Cancerome Analysis Network–Breast; ER = estrogen receptor; PR = progesterone receptor; HER2 = human epidermal growth factor receptor 2.

|  | MyBrCa (%) |  | SCAN-B (%) | METABRIC (%) |
| --- | --- | --- | --- | --- |
|  | PREDICT validation | Machine learning development |  |  |
| **Total** | 3875 | 258 | 2072 | 758 |
| **Median follow-up (years)** | 7.3 | 8.6 | 7.5 | 11.0 |
| **Deaths (n)** | 709 (18.3%) | 134 (51.9%) | 1188 (57.3%) | 490 (64.6%) |
| **Median age at diagnosis** | 51 | 53 | 70 | 61 |
| **Age group** |  |  |  |  |
| < 36 y | 238 (6.1%) | 15 (5.8%) | 20 (1.0%) | 23 (3.0%) |
| 36-40 y | 294 (7.6%) | 16 (6.2%) | 33 (1.6%) | 21 (2.8%) |
| 41-50 y | 1298 (33.5%) | 78 (30.2%) | 234 (11.3%) | 128 (16.9%) |
| 51-60 y | 1162 (30.0%) | 70 (27.1%) | 313 (15.1%) | 170 (22.4%) |
| > 60 y | 883 (22.8%) | 79 (30.6%) | 1472 (71.0%) | 416 (54.9%) |
| **ER/PR/HER2 status** |  |  |  |  |
| ER+ | 2708 (69.9%) | 150 (58.1%) | 1673 (80.7%) | 601 (79.3%) |
| PR+ | 2390 (61.7%) | 120 (46.5%) | 1336 (64.5%) | 389 (51.3%) |
| HER2+ | 924 (23.8%) | 79 (30.6%) | 279 (13.5%) | 94 (12.4%) |
| **Stage** |  |  |  |  |
| 0 | 136 (3.5%) | 0 (0.0%) | 0 (0.0%) | 1 (0.1%) |
| 1 | 1277 (33.0%) | 35 (13.6%) | 1145 (55.3%) | 256 (33.8%) |
| 2 | 1698 (43.8%) | 120 (46.5%) | 842 (40.6%) | 438 (57.8%) |
| 3 | 764 (19.7%) | 103 (39.9%) | 85 (4.1%) | 63 (8.3%) |
| **Grade** |  |  |  |  |
| 0 | 1 (0.0%) | 0 (0.0%) | 0 (0.0%) | 0 (0.0%) |
| 1 | 385 (9.9%) | 4 (1.6%) | 250 (12.1%) | 56 (7.4%) |
| 2 | 2020 (52.1%) | 111 (43.0%) | 962 (46.4%) | 302 (39.8%) |
| 3 | 1469 (37.9%) | 143 (55.4%) | 860 (41.5%) | 400 (52.8%) |
| **Size (cm)** |  |  |  |  |
| ≤1 | 439 (11.3%) | 11 (4.3%) | 222 (10.7%) | 21 (2.8%) |
| >1, ≤ 2 | 1323 (34.1%) | 49 (19.0%) | 923 (44.5%) | 308 (40.6%) |
| >2, ≤ 5 | 1785 (46.1%) | 168 (65.1%) | 842 (40.6%) | 396 (52.2%) |
| >5 | 328 (8.5%) | 30 (11.6%) | 85 (4.1%) | 33 (4.4%) |
| **Positives nodes** |  |  |  |  |
| 0 | 2397 (61.9%) | 110 (42.6%) | 1243 (60.0%) | 387 (51.1%) |
| >1, ≤ 3 | 796 (20.5%) | 55 (21.3%) | 520 (25.1%) | 245 (32.3%) |
| >3, ≤ 9 | 408 (10.5%) | 43 (16.7%) | 309 (14.9%) | 84 (11.1%) |
| ≥ 10 | 274 (7.1%) | 50 (19.4%) | 0 (0.0%) | 42 (5.5%) |
| **Adjuvant chemotherapy** | 396 (10.2%) | 100 (38.8%) | 0 (0.0%) | 0 (0.0%) |
| **Trastuzumab** | 82 (2.1%) | 112 (43.4%) | 0 (0.0%) | 0 (0.0%) |
| **Adjuvant hormone therapy** | 428 (11.0%) | 24 (9.3%) | 1553 (75.0%) | 460 (60.7%) |
| **Radiation therapy** | 395 (10.2%) | 107 (41.5%) | 0 (0.0%) | 506 (66.8%) |
| **Classification** |  |  |  |  |
| Poor | NA | 124 (48.1%) | 1178 (56.9%) | 330 (43.5%) |
| Good | NA | 134 (51.9%) | 894 (43.1%) | 428 (56.5%) |

The largest proportion of patients in this cohort was classified as having grade 2 tumors (moderately differentiated; 52.1%), followed by grade 3 (poorly differentiated; 37.9%) and grade 1 (well-differentiated; 9.9%). Although treatment data are optional inputs for the PREDICT model, they were incomplete for most of the patients. Only a limited proportion of patients (10.2% to 11.0%) had available treatment information for adjuvant chemotherapy, adjuvant hormone therapy, or radiation therapy.

From this dataset, a subset of 258 patients was selected for prognostic model development (Table 1). The selection was based on the availability of RNA-seq data and complete HER2 and PR receptor status. Only patients who could be classified into good or poor prognosis groups were included. Applying the same exclusion and classification criteria, 2072 patients from the SCAN-B cohort and 758 patients from the METABRIC cohort were selected as external validation cohorts. Notably, most of the patients in the external cohorts were excluded because they were still alive but had not yet reached 10 years of follow-up; therefore, could not be reliably assigned a prognosis group. The METABRIC cohort has the highest median follow-up (11 years) and recorded the highest proportion of deaths (64.6%). The cohorts also differed in age at diagnosis, with MyBrCa being younger at diagnosis (median: 53 years) and SCAN-B being the oldest (median: 70 years). This was reflected in the proportion of patients over 60 at diagnosis, which was the highest in SCAN-B (71.0%) compared to 54.9% in METABRIC and 30.6% in MyBrCa. ER and PR positivity were the highest in SCAN-B, suggesting a larger proportion of hormone receptor-positive tumors, which are generally less aggressive.

Consistent with this, a higher proportion of patients in SCAN-B were diagnosed with early stage of disease (55.3% at stage 1 and 40.6% at stage 2) in comparison to METABRIC (33.8% and 57.8%) and MyBrCa (13.6% and 46.5%). Taken together, these differences highlight that the MyBrCa dataset used for model development represents a younger cohort with higher proportion of patients with advanced disease, SCAN-B reflects an older population with higher proportion of hormone receptor–positive and early-stage tumor, while METABRIC being in between with intermediate age at diagnosis and the highest proportion of deaths due to its longer follow-up. Despite these differences, relatively balanced datasets were obtained when these datasets were stratified using a 10-year survival cut-off. This suggests the suitability of the 10-year threshold for model development and evaluation.

### 4.2 Validation of PREDICT v3.0 in the MyBrCa cohort

The 5-,10-, and 15-year overall survival estimates by the Kaplan-Meier (KM) analysis was 89.6%, 79.2%, and 70.8% with a corresponding 95% CI of 88.6%-90.6%, 77.6%-80.8%, and 68.5%-73.3%, respectively (Table 2). In comparison, PREDICT v3.0 estimated higher median survival probabilities of 94.2%, 85.7%, and 77.6% with IQRs of 86.4%-97.2%, 75.4%-92.6%, and 65.0%-86.9%, respectively. The corresponding AUROCs were consistent across the 5-, 10-, and 15- year predictions at approximately 0.69 (95% CI: 0.67–0.72) (Figure 2A).

**Figure 2.**
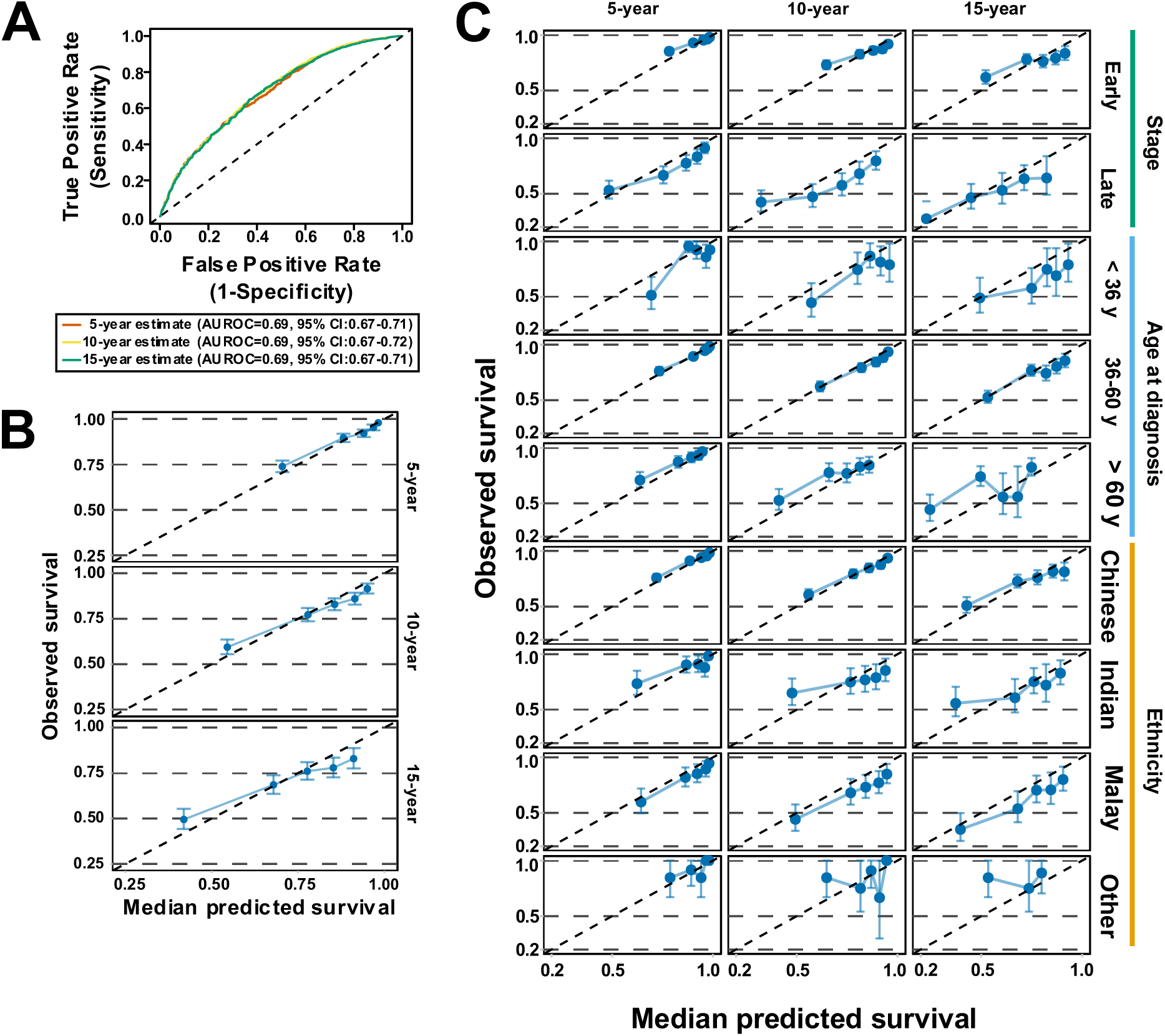
Evaluation of PREDICT’s performance in the MyBrCa cohort. **A** Receiver operating characteristic (ROC) curve showing the discriminative performance of PREDICT v3.0 for estimating survival at three timepoints (5,10, and 15 years) for 3875 MyBrCa patients. Model performance is quantified by the AUROC. **B** Calibration plot comparing mean predicted versus observed survival in the full MyBrCa cohort. Predictions were binned into quintiles for visualization. The diagonal line represents perfect calibration performance. For clarity, both axes are cropped to begin at 0.25. **C** Calibration plots visualizing PREDICT’s calibration in selected subgroups of patients in the MyBrCa cohort. Axes are cropped to begin at 0.2.

**Table 2.** Comparison of observed 5-, 10-, and 15-year overall survival (OS) with survival probabilities estimated by PREDICT v3.0. Observed OS based on Kaplan–Meier estimates were 89.6 (95% CI: 88.6-90.6), 79.2 (95% CI: 77.6-80.8), and 70.8 (95% CI: 68.5-73.3) at 5, 10, and 15 years, respectively. In contrast, the median PREDICT-estimated survival probabilities for the same time points were 94.2 (IQR: 86.4-97.2), 85.7 (IQR: 75.4-92.6), and 77.6 (IQR: 65-86.9). Results were further stratified by age group, ethnicity, tumor stage, grade, clinical subtype, receptor status, and systemic therapy. Values for 15-year OS in the systemic therapy category were reported as NA due to the absence of patients with 15-year follow-up. Δ% (percent difference) represents the relative difference between KM-derived overall and PREDICT-estimated survival, calculated as [(Observed − Predicted) / Observed] × 100. IQR = interquartile range; CI = confidence interval.

|  | n | 5-year survival |  |  | 10-year survival |  |  | 15-year survival |  |  |
| --- | --- | --- | --- | --- | --- | --- | --- | --- | --- | --- |
|  |  | Overall survival<br>(95% CI) | PREDICT-estimated<br>median survival<br>(IQR) | Δ<br>(%) | Overall survival<br>(95% CI) | PREDICT-estimated<br>median survival<br>(IQR) | Δ<br>(%) | Overall survival<br>(95% CI) | PREDICT-estimated<br>median survival<br>(IQR) | Δ<br>(%) |
| **All patients** | 3875 | 89.6 (88.6-90.6) | 94.2 (86.4-97.2) | **4.9** | 79.2 (77.6-80.8) | 85.7 (75.4-92.6) | **7.6** | 70.8 (68.5-73.3) | 77.6 (65-86.9) | **8.8** |
| **Age** |  |  |  |  |  |  |  |  |  |  |
| < 36 y | 238 | 83.2 (78.4-88.3) | 91.2 (86.9-97.3) | **8.8** | 72.9 (66.7-79.7) | 86.4 (77.6-93) | **15.6** | 65.2 (57.7-73.8) | 82.7 (71.9-88.6) | **21.2** |
| 36-40 y | 294 | 91.8 (88.6-95.2) | 96.9 (89.6-98.4) | **5.3** | 82.2 (77-87.7) | 91.9 (83.6-95.6) | **10.6** | 72.3 (64.3-81.2) | 86.7 (79.3-92.1) | **16.6** |
| 41-50 y | 1298 | 90.5 (88.8-92.2) | 97.1 (89.3-98) | **6.8** | 80.9 (78.3-83.5) | 92.4 (82.3-94.6) | **12.4** | 75.2 (71.6-79) | 86.7 (76.2-90.2) | **13.3** |
| 51-60 y | 1162 | 90.8 (89.1-92.6) | 95.1 (86.5-96.7) | **4.5** | 80.9 (78.2-83.7) | 87.2 (77.1-91) | **7.2** | 71.6 (67.4-76.1) | 78.3 (69.1-83.7) | **8.6** |
| > 60 y | 883 | 87.7 (85.4-90.1) | 90 (81.3-93.5) | **2.6** | 74.7 (71.1-78.6) | 75.1 (63.6-82.7) | **0.5** | 61.9 (55.1-69.5) | 60.8 (45.4-69.8) | **-1.8** |
| **Self-reported ethnicity** |  |  |  |  |  |  |  |  |  |  |
| Chinese | 3038 | 90.8 (89.7-91.9) | 94.4 (86.9-97.3) | **3.8** | 81 (79.2-82.8) | 85.9 (75.7-92.8) | **5.7** | 72.6 (69.7-75.6) | 78 (65.3-87.2) | **6.9** |
| Indian | 322 | 88.3 (84.7-91.9) | 92.9 (85.2-96.5) | **5.0** | 76.4 (71.5-81.6) | 84.1 (74.2-90.5) | **9.2** | 69.2 (63.2-75.7) | 76.3 (63.7-83.7) | **9.3** |
| Malay | 394 | 82.3 (78.5-86.2) | 92.4 (84.7-97) | **10.9** | 69.6 (64.9-74.6) | 84.3 (74.3-92) | **17.4** | 62.1 (56.5-68.2) | 77.4 (65.3-86) | **19.8** |
| Other | 63 | 91.9 (85.3-99) | 94.8 (88.2-97) | **3.1** | 85.5 (75.3-97) | 86.7 (80.4-91.9) | **1.4** | 68.4 (43.3-100) | 80 (70.9-86.1) | **14.5** |
| Unknown | 55 | 79.4 (64.4-97.8) | 93.6 (81.8-97.5) | **15.2** | 74.1 (57.8-95) | 84.6 (69.6-93.1) | **12.4** | NA | 73.7 (59.7-87.5) | **NA** |
| **Stage** |  |  |  |  |  |  |  |  |  |  |
| Early stage (0-II) | 3111 | 93.5 (92.6-94.4) | 95.4 (88.5-97.5) | **2.0** | 84.1 (82.5-85.7) | 88 (79.5-93.3) | **4.4** | 75.6 (73.1-78.3) | 80.7 (70.1-88.1) | **6.3** |
| Late stage (III) | 764 | 74.1 (70.9-77.4) | 86.9 (71.3-93) | **14.7** | 58.7 (54.4-63.3) | 72.9 (53.4-83) | **19.5** | 49.6 (43.4-56.7) | 60.5 (41-73.8) | **18.0** |
| **Grade** |  |  |  |  |  |  |  |  |  |  |
| Grade I | 385 | 98.1 (96.7-99.6) | 97.5 (95.2-98.3) | **-0.6** | 92.1 (88.8-95.6) | 93.2 (87.9-95.3) | **1.2** | 77.2 (70.2-85) | 87.9 (79.5-91.3) | **12.2** |
| Grade II | 2020 | 92.2 (90.9-93.5) | 95.7 (90.4-97.6) | **3.7** | 79.9 (77.6-82.2) | 88.6 (79.4-93.5) | **9.8** | 72.1 (68.7-75.6) | 80.5 (68.7-88.4) | **10.4** |
| Grade III | 1469 | 84.1 (82.2-86) | 88.2 (81.3-94.4) | **4.6** | 74.7 (72.1-77.3) | 79.5 (68.5-86.5) | **6.0** | 67.7 (64.1-71.5) | 71.8 (57.3-80.6) | **5.7** |
| **Clinical subtype** |  |  |  |  |  |  |  |  |  |  |
| TNBC | 566 | 81.5 (78.2-84.8) | 85.7 (77.8-88.9) | **4.9** | 73.9 (70-78.1) | 77.5 (65.8-83.1) | **4.6** | 67.2 (62.1-72.8) | 71.4 (58-78.9) | **5.9** |
| HER2Pos | 464 | 82.7 (79.3-86.4) | 80 (66.3-84.6) | **-3.4** | 73.5 (69.1-78.2) | 70.3 (53.2-76.7) | **-4.6** | 67.9 (62.5-73.8) | 62.6 (45.7-71.4) | **-8.5** |
| HRPosHER2Neg | 1981 | 93.4 (92.2-94.6) | 96.5 (93.6-97.8) | **3.2** | 81.3 (79-83.7) | 90.7 (83.4-94.1) | **10.4** | 71.4 (67-76) | 83.6 (71.7-89.5) | **14.6** |
| HRPosHER2Pos | 454 | 89.8 (86.9-92.7) | 95.5 (92.4-97.3) | **6.0** | 80.6 (76.4-85) | 88.2 (80.8-92.7) | **8.6** | 71.6 (65.4-78.4) | 79.8 (68.8-86.9) | **10.3** |
| Unclassified | 410 | 91.1 (88.3-94.1) | 94.7 (87.4-97.1) | **3.8** | 80 (75.8-84.5) | 86.2 (77.3-92.1) | **7.2** | 70.9 (65.2-77.1) | 78.7 (68.3-86.3) | **9.9** |
| **ER status** |  |  |  |  |  |  |  |  |  |  |
| ER-negative | 1167 | 82.3 (80-84.5) | 83.4 (73-87.7) | **1.3** | 74.6 (71.8-77.4) | 74.4 (60.8-81) | **-0.3** | 68.8 (65.3-72.4) | 67.5 (52.5-75.8) | **-1.9** |
| ER-positive | 2708 | 92.9 (91.9-94) | 96.4 (93.6-97.7) | **3.6** | 81.1 (79.1-83) | 90.3 (83.2-93.8) | **10.2** | 71 (67.8-74.3) | 82.9 (71.5-89) | **14.4** |
| **PR status** |  |  |  |  |  |  |  |  |  |  |
| PR-negative | 1465 | 84.6 (82.7-86.5) | 85.3 (77.2-90) | **0.8** | 76.2 (73.7-78.8) | 76.4 (64.2-83.4) | **0.3** | 69.4 (66-73.1) | 69.2 (54.5-77.6) | **-0.3** |
| PR-positive | 2390 | 92.8 (91.7-93.9) | 96.5 (93.7-97.8) | **3.8** | 81.3 (79.3-83.3) | 90.7 (83.8-94) | **10.4** | 71.8 (68.6-75.1) | 83.8 (72.4-89.3) | **14.3** |
| Unknown | 20 | 85 (70.7-100) | 90.5 (81.1-96.5) | **6.1** | 59.1 (40.7-85.7) | 81.6 (73.6-90.9) | **27.6** | 51.7 (32.8-81.5) | 73.7 (67.5-84.8) | **29.9** |
| **HER2 status** |  |  |  |  |  |  |  |  |  |  |
| HER2-negative | 2548 | 90.6 (89.4-91.9) | 95.1 (89.2-97.5) | **4.7** | 79.8 (77.8-81.8) | 87.4 (79-93.3) | **8.7** | 71 (67.7-74.5) | 80.2 (68.2-88.2) | **11.5** |
| HER2-positive | 924 | 86.3 (84.1-88.6) | 87 (78.9-95.5) | **0.8** | 76.8 (73.7-80) | 78.1 (65.3-88.3) | **1.7** | 69.3 (65.1-73.7) | 70.3 (54.8-80.7) | **1.4** |
| Unknown | 403 | 91 (88.1-94) | 94.7 (87.5-97.1) | **3.9** | 80.3 (76-84.8) | 86.3 (77.4-92.2) | **7.0** | 71.4 (65.7-77.6) | 78.7 (68.3-86.4) | **9.3** |
| **Systemic therapy** |  |  |  |  |  |  |  |  |  |  |
| No chemotherapy or hormone therapy | 212 | 76.4 (70.9-82.3) | 84.6 (71.6-93.8) | **9.7** | 62.1 (54.9-70.3) | 74.3 (53.8-85.8) | **16.4** | NA | 64.7 (43.4-78.6) | **NA** |
| Chemotherapy and hormone therapy | 245 | 94.3 (91.4-97.2) | 96.2 (93.6-97.4) | **2.0** | 79.6 (73.4-86.2) | 89.6 (83.2-93.1) | **11.2** | NA | 81.7 (72-87.7) | **NA** |
| Chemotherapy only | 180 | 86.1 (81.2-91.3) | 87.6 (83.1-91.2) | **1.7** | 82.1 (76.4-88.3) | 79.2 (71.7-85.1) | **-3.7** | NA | 72.2 (60.4-79.2) | **NA** |
| Hormone therapy only | 183 | 89.1 (84.7-93.7) | 95.6 (91.6-97.8) | **6.8** | 77.6 (69.9-86.2) | 88.4 (78.3-94) | **12.2** | NA | 79.6 (62.2-89.2) | **NA** |
| Trastuzumab | 82 | 92.7 (87.2-98.5) | 91.9 (85.6-96.1) | **-0.9** | 90.5 (83.9-97.7) | 84.9 (73-90.2) | **-6.6** | NA | 76.7 (60.8-85.7) | **NA** |
| Radiation therapy | 395 | 88.1 (85-91.4) | 93.5 (86.9-97) | **5.8** | 76.7 (71.7-82) | 85.2 (76-92) | **10.0** | NA | 76.9 (63.5-86.3) | **NA** |

When comparing KM-derived overall survival with PREDICT-estimated median survival, PREDICT v3.0 showed good calibration overall, with percent differences not exceeding 10% of the KM-derived values (Table 2). However, across demographics subgroups, poor calibration was observed for specific age subgroups (<36 years, 36–40 years, and 41–50 years) at long-term predictions (10- and 15-year) and across all timepoints for patients of Malay ethnicity. Across clinical subgroups, PREDICT v3.0 demonstrated poor calibration at long-term predictions in hormone receptor–positive (HR+) subtypes, including ER- and PR-positive disease, as well as in HER2-negative disease. Notably, the calibration was poor all timepoints for patients with late-stage cancer (Stage 3). For patients with known treatment information, PREDICT v3.0 predictions were within 10% of KM-derived survival at 5 years, but showed poor calibration at 10 years in patients receiving no systemic therapy, chemotherapy plus hormone therapy, or hormone therapy alone. Notably, predictions were accurate at both time points in three distinct subgroups: patients receiving hormone therapy, patients receiving trastuzumab, and patients receiving radiation therapy.

Goodness-of-fit assessment revealed that PREDICT v3.0 overestimated survival in the lowest-risk quintile and underestimated survival in the highest-risk quintile, with deviations more pronounced for long-term 10- and 15-year predictions (Figure 2B). Across demographic and clinical subgroups, poor calibration in higher-risk quintiles was particularly evident among patients with late-stage cancer (Stage 3), patients diagnosed under 36 years, patients diagnosed over 60 years, patients of Malay ethnicity (noting the small sample size in the ‘Other’ ethnic group), and in patients with HER2-positive disease (Figure 2C, Supplementary Figure 2). The predictions were closer to observed survival in most quintiles for with triple-negative breast cancer (TNBC) and in patients with ER- and PR-positive disease. For patients with known treatment information, overall survival was generally high, and PREDICT v3.0 estimates reflected this trend, with median survival values closely matching those observed. However, it still underestimated the 10-year survival in patients who received trastuzumab.

Taken together, these findings suggest that while PREDICT v3.0 demonstrated moderate discriminative performance, its estimates were poorly calibrated for long-term predictions and specific patient subgroups, particularly those at higher risk, highlighting the potential for misinformed clinical decisions in vulnerable populations.

### 4.3 Selection of prognostic gene signature in the MyBrCa cohort

The selected 258 MyBrCa patients’ global transcriptome dataset comprised 18,112 cleaned expressed genes. The principal component analysis revealed no distinct clustering by clustering, indicating a high degree of overlap in the transcriptomic profiles of the MyBrCa patients in the two prognosis groups (Supplementary Figure 3A). After retaining the top 30 genes with the highest loading coefficient from each of the top principal components that cumulatively account for 80% of the total variance (94 PCs), a reduced list of 995 genes was obtained. Since LASSO was applied in the subsequent step, genes were retained from the PCs to allow regularization to operate on a broader set of candidate genes, including those associated with lower variance but potentially prognostic. The cut-off of 30 was selected based on an ’elbow point’ observed during comparisons with other threshold values (Supplementary Figure 3B); beyond this threshold, including additional genes yielded minimal gains in the variance captured by each principal component. The final gene list was fitted in a LASSO regression model for multicollinearity reduction (Supplementary Figure 3C). The optimal value of the hyperparameter lambda, which dictates the strength of penalization applied to the coefficients in the model, was identified as 0.026 using the AIC. In comparison, the BIC, which enforces a stricter penalization than AIC, selected the optimal value of 0.11 (Figure 3A). The final model, consisting of 77 genes with non-zero coefficients, was obtained using the AIC-derived lambda (Figure 3B). From this point onwards, this gene signature of 77 genes is referred to as MyGESig.

**Figure 3.**
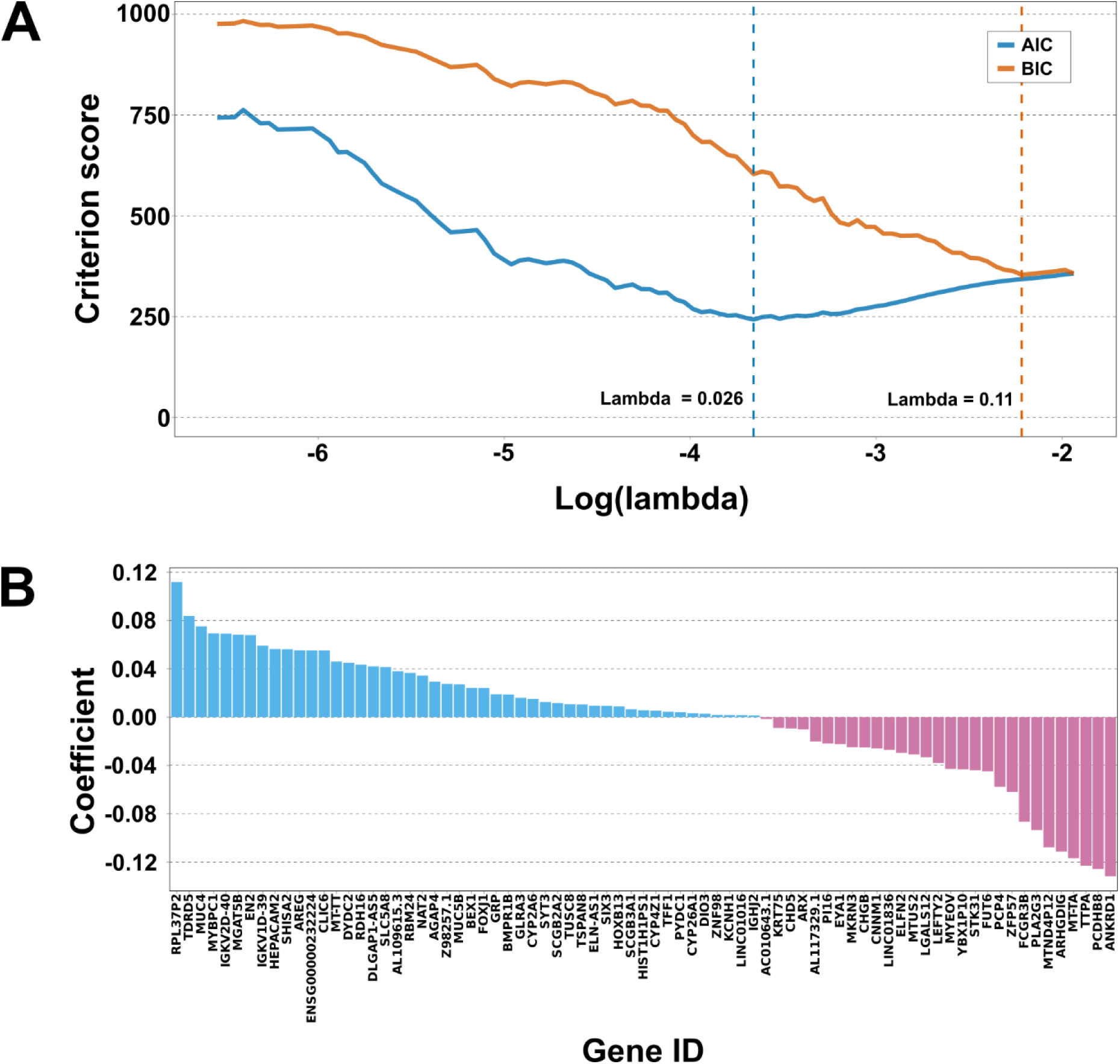
AIC/BIC-guided lambda selection and retained genes in LASSO. **A** Plot of AIC and BIC scores across a range of log-transformed lambda values. The optimal lambda was selected based on the minimum values of both criteria. **B** Bar plot summarizing the non-zero gene coefficients retained in the final LASSO model.

GO term classification revealed that these genes were associated with regulation and signalling (GO:0050794, regulation of cellular process; GO:0023052, signalling; GO:0007165, signal transduction), development and differentiation (GO:0007275, multicellular organism development, GO:0030154, cell differentiation), biosynthesis (GO:0044249, cellular biosynthetic process; GO:1901576, organic substance biosynthetic process), metabolism (GO:0043170, macromolecule metabolic process), and cellular organization (GO:0016043, cellular component organization) (Supplementary Figure 4). On the other hand, classification of molecular function categories revealed the association of these genes with functions related to binding (GO:0043169, cation binding; GO:0043168, anion binding; GO:0003676, nucleic acid binding; GO:0005102, signalling receptor binding), transcriptional regulation (GO:0000981, DNA-binding transcription factor activity, RNA polymerase II-specific), receptor activation (GO:0030546, signalling receptor activator activity), and transporter activities (GO:0015318, inorganic molecular entity transmembrane transporter activity; GO:0022803, passive transmembrane transporter activity; GO:1901702, salt transmembrane transporter activity; GO:0015075, monoatomic ion transmembrane transporter activity) (Supplementary Figure 4). Subsequent gene set enrichment analysis using the hallmark gene sets (H) from the MSigDB returned no statistically significant enrichment after multiple testing correction (adjusted p-value < 0.05). Subsequent gene set enrichment analysis using the hallmark gene sets (H) from the MSigDB returned no statistically significant enrichment after multiple testing correction (adjusted p-value < 0.05) (Supplementary Table 1). This indicates that this gene signature is not associated with the hallmark biological pathways represented in the MSigDB.

### 4.4 Classification of prognosis using the MyGESig gene signature

A prognostic model was developed to use MyGESig gene signature and clinical information (age at diagnosis, hormone receptor (ER/PR) status, HER2 status, tumor grade, tumor size, and number of locoregional lymph nodes to which cancer has spread) to classify the MyBrCa patients based on their 10-year prognosis (Supplementary Figure 1). The clinical information was selected based on the key inputs used by the PREDICT v3.0 tool, excluding treatment information and smoking status due to limited data availability. The implementation of this model is based on a classifier described in Sammut et. al. (2020). This adaptation involves a 5- seed stratified split, allocating 70% of the patients for model training and 30% for testing. 5-fold cross-validation was utilized to evaluate the model’s performance to ensure robustness and minimal variability across different training subsets. This classifier trained using both clinical variables and the MyGESig signature is hereafter referred to as the MyBrCa70-Combined model (i.e., trained on 70% of the MyBrCa dataset with both clinical and gene features). For clarity, MyBrCa70 is used as the base model name, with suffixes (e.g., -Clinical, -MyGESig, -Combined) denoting the specific feature set used for training. Utilizing both gene expression and clinical data as features, the MyBrCa70-Combined model achieved a median AUROC of 0.99 (95% CI: 0.98–0.99) and 0.92 (95% CI: 0.87–0.92) across five training and five testing datasets, respectively (Figure 4A, Supplementary Table 2). Using the probability threshold that maximizes the Youden Index, the model achieved the accuracy, specificity, and sensitivity of 0.88, 0.95, and 0.81, in the testing dataset, respectively (Table 3). Furthermore, the individual algorithms in this model exhibited strong discriminative performance with the AUROC of 0.83 (95% CI: 0.74– 0.83) (random forest) and 0.92 (95% CI: 0.87–0.92) (support vector machine) (Figure 4B, Supplementary Table 2).

**Figure 4.**
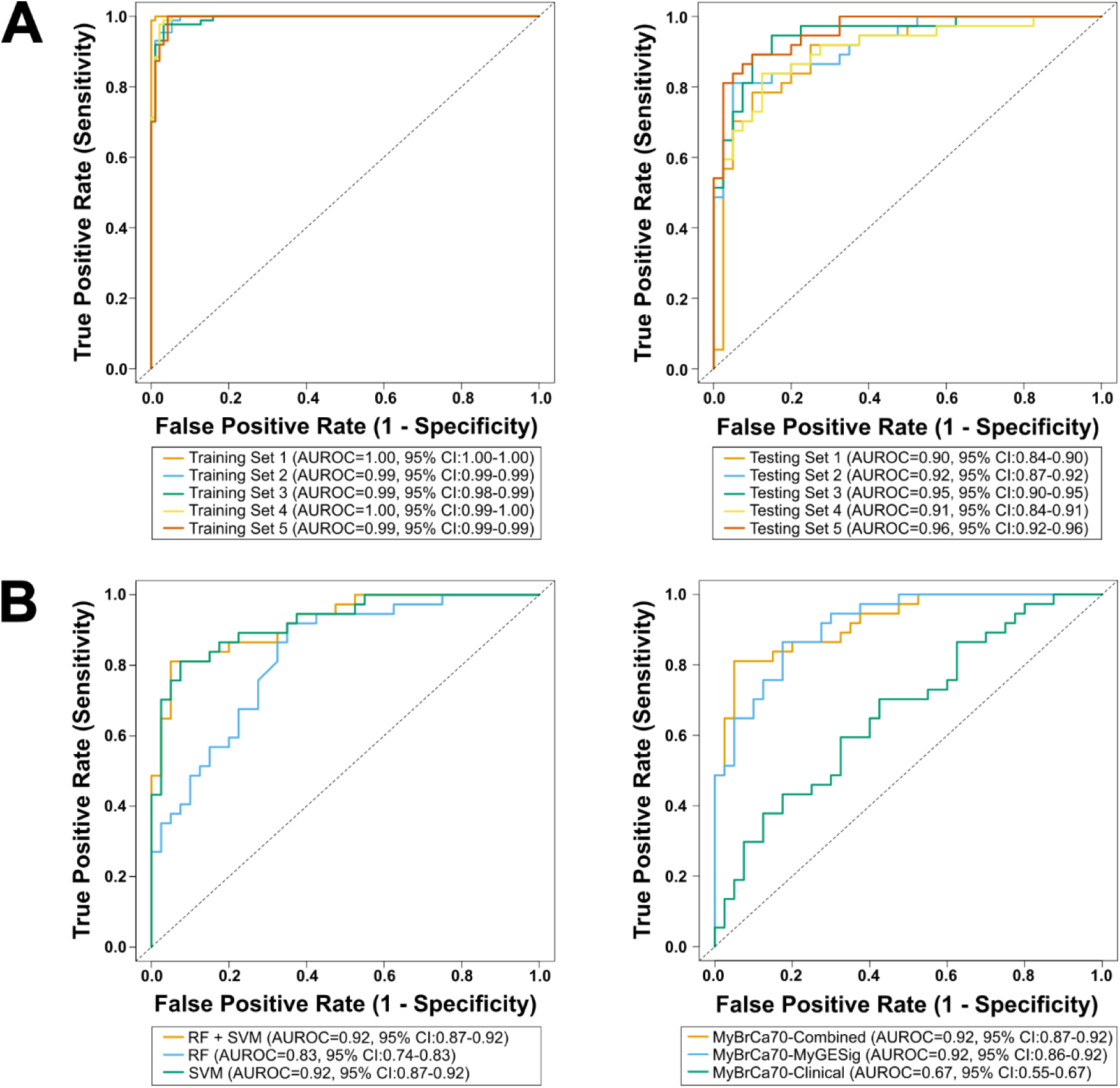
Evaluation of model discriminative performance across different datasets, algorithms and feature sets. **A** ROC curves showing model performance in classifying MyBrCa patients according to their 10-year prognosis in the 70:30 split for training (left) and testing (right) datasets. The model was trained using the LASSO-derived 77-gene signature, termed MyGESig, combined with clinical information (referred to as MyBrCa70-Combined model). **B** ROC curves comparing model performance when using different machine learning algorithms (left) and when trained on different combinations of features (right).

**Table 3.** Comparison of machine learning model performance trained on 70% of the MyBrCa dataset using different combination of features. Performance metrics were calculated using a probability threshold that maximizes the Youden Index. The performance of the MyGESig gene signature was compared with established Western-based (Mammaprint, Oncotype DX, and PAM50) and the Eastern-based signatures (GenesWell BCT, signature from Cheng et. al. (2016)). AUROC = area under the receiver-operator characteristic curve.

|  | **AUROC (95% CI)** | **Accuracy** | **Kappa** | **Sensitivity** | **Specificity** | **Precision** | **F1 score** |
| --- | --- | --- | --- | --- | --- | --- | --- |
| **Clinical information** | 0.67 (0.55-0.67) | 0.64 | 0.28 | 0.70 | 0.58 | 0.60 | 0.65 |
| **Gene expression** |  |  |  |  |  |  |  |
| MyGESig | 0.92 (0.86-0.92) | 0.84 | 0.69 | 0.86 | 0.83 | 0.82 | 0.84 |
| Oncotype DX | 0.60 (0.47-0.60) | 0.65 | 0.31 | 0.89 | 0.43 | 0.59 | 0.71 |
| PAM50 | 0.61 (0.49-0.61) | 0.62 | 0.23 | 0.27 | 0.95 | 0.83 | 0.41 |
| MammaPrint | 0.64 (0.52-0.64) | 0.61 | 0.24 | 0.92 | 0.33 | 0.56 | 0.69 |
| GenesWell BCT | 0.54 (0.41-0.54) | 0.43 | -0.14 | 0.49 | 0.38 | 0.42 | 0.45 |
| Cheng et. al. (2016) | 0.59 (0.46-0.59) | 0.66 | 0.31 | 0.46 | 0.85 | 0.74 | 0.57 |
| **Gene expression + Clinical information** |  |  |  |  |  |  |  |
| MyGESig | 0.92 (0.87-0.92) | 0.88 | 0.76 | 0.81 | 0.95 | 0.94 | 0.87 |
| Oncotype DX | 0.73 (0.61-0.73) | 0.70 | 0.41 | 0.81 | 0.60 | 0.65 | 0.72 |
| PAM50 | 0.69 (0.58-0.69) | 0.66 | 0.33 | 0.73 | 0.60 | 0.63 | 0.68 |
| MammaPrint | 0.71 (0.59-0.71) | 0.70 | 0.40 | 0.76 | 0.65 | 0.67 | 0.71 |
| GenesWell BCT | 0.65 (0.52-0.65) | 0.65 | 0.28 | 0.32 | 0.95 | 0.86 | 0.47 |
| Cheng et. al. (2016) | 0.68 (0.56-0.68) | 0.65 | 0.28 | 0.32 | 0.95 | 0.86 | 0.47 |

The contribution and complementarity of clinical data and MyGESig signature in the MyBrCa70-Combined model was assessed by retraining it with either feature set alone. When trained solely on clinical data (hence referred to as the MyBrCa70-Clinical model), the resulting model achieved a median AUROC of 0.67 (95% CI: 0.55–0.69) (Figure 4B, Table 3), which was marginally lower than PREDICT v3.0’s 10-year survival prediction (AUROC = 0.69). Notably, the clinical variables used for training were the same as those required by the PREDICT model, and both models were validated in datasets that differed markedly in sample size. On the other hand, when trained solely on the MyGESig signature (MyBrCa70-MyGESig model), it demonstrated a stronger discriminative performance, achieving a median AUROC of 0.92 (95% CI: 0.86–0.92). This indicates that transcriptome-derived molecular signals captured by this signature are better at distinguishing between good and poor prognosis groups in the MyBrCa cohort, compared to the clinical variables. When established gene signatures (Mammaprint, Oncotype DX, and PAM50) were used to predict prognosis in the MyBrCa cohort, the discriminative performance dropped significantly, with the median AUROCs of 0.64, 0.60, and 0.61, respectively (95% CIs: 0.52–0.64, 0.47–0.60, and 0.49–0.61) (Table 3, Supplementary Figure 5A). The Eastern-based gene signatures were also tested [GenesWell BCT and the 34- gene signature from Cheng et al. (2016)], considering that most established signatures are Western-centric, to further evaluate performance in the MyBrCa cohort, achieving median AUROCs of 0.54 (95% CIs: 0.41-0.54) and 0.59 (95% CIs: 0.46-0.59). The poor performance of these gene signatures underlines the limitation of these signatures to capture strong prognosis-related variance in the transcriptome of the MyBrCa cohort.

As a means of validating the feature selection approach, the shrinkage of less informative genes was assessed by iterative LASSO modelling was performed across a fine-grained sequence of decreasing lambda values, starting from the AIC-derived value (lambda = 0.026). During each iteration, genes with non-zero coefficients were identified and contrasted against the genes identified in the subsequent iteration, retaining only genes uniquely kept at that specific lambda. For a fair comparison to the 77-gene MyGESig, only gene sets with at least 70 non-overlapping genes were considered. When models were trained using the respective gene sets, performance improved with increasing lambda, indicating that stronger penalization isolates genes that are more informative and can discriminate prognosis better, as reflected in improved AUROC metrics (Figure 5).

**Figure 5.**
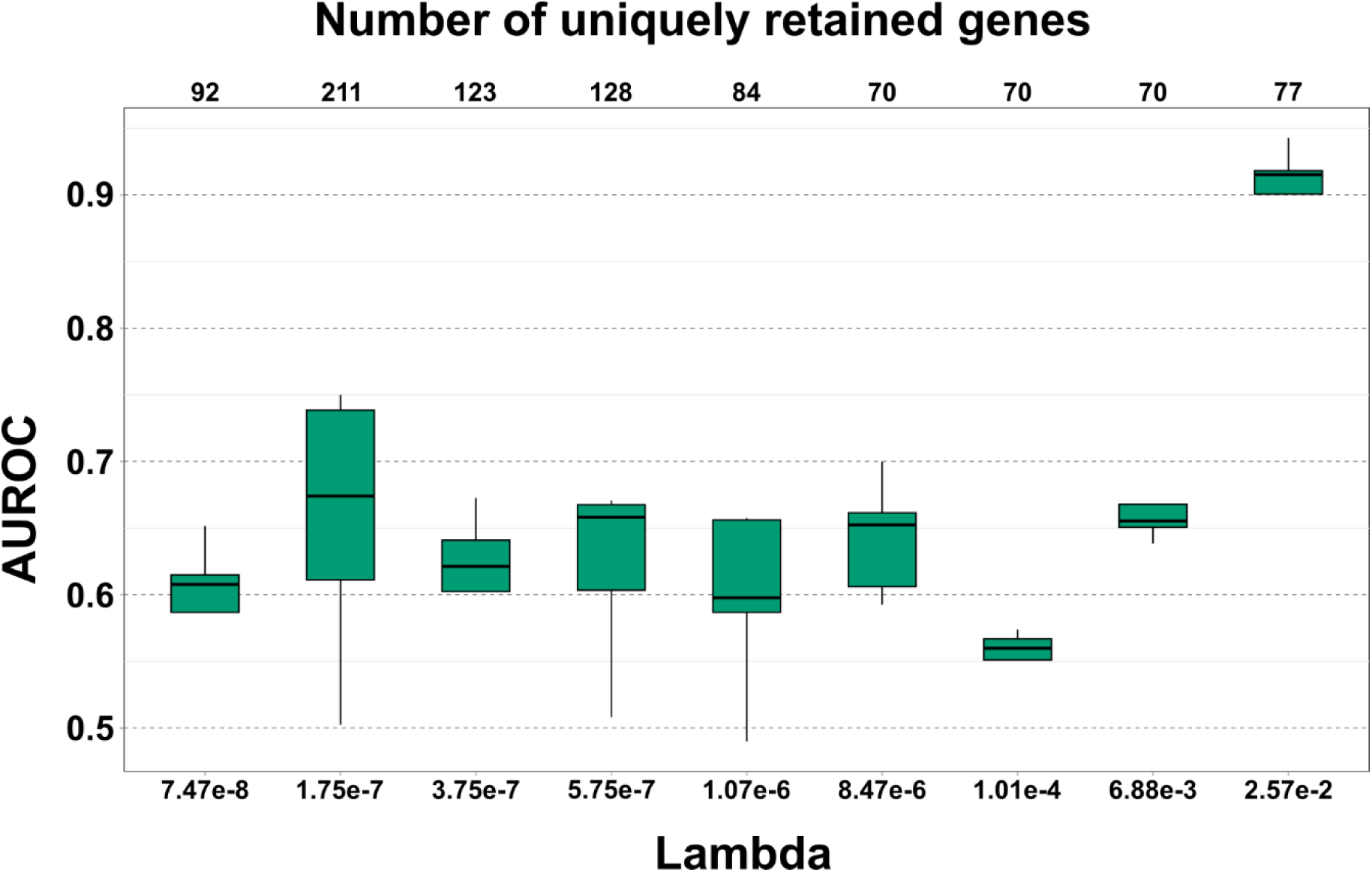
Discriminative performance of uniquely retained genes across varying regularization strengths. Box-and-whisker plots summarizing the AUROC scores of machine learning models trained using uniquely retained gene sets derived from LASSO regression at different levels of penalization (represented by lambda). Gene sets retained at higher lambda values represent more strongly penalized, and potentially more informative, subsets. The gene set at lambda = 2.57e-2 corresponds to the MyGESig signature.

Considering the MyBrCa70 models, namely the MyGESig and Combined variants, were trained using the 77 genes identified from the entire machine learning subset (n = 258), there could be information leakage, potentially leading to overfitting. To rule this out, the models were retrained on a reduced subset of training data and evaluated in the independent MyBrCa dataset by applying the stratified split twice and retaining the first 30% split as an independent dataset (Supplementary Figure 1). When the resulting dataset of 77 patients was used for validation, the newly derived models (hereafter referred to as the IndMyBrCa30 models) achieved median AUROCs of 0.74, 0.84, and 0.90 when using clinical data, gene signatures, or both, respectively (95% CIs: 0.63–0.74, 0.76–0.84, and 0.83–0.90) (Figure 6A, Supplementary Table 2).

**Figure 6.**
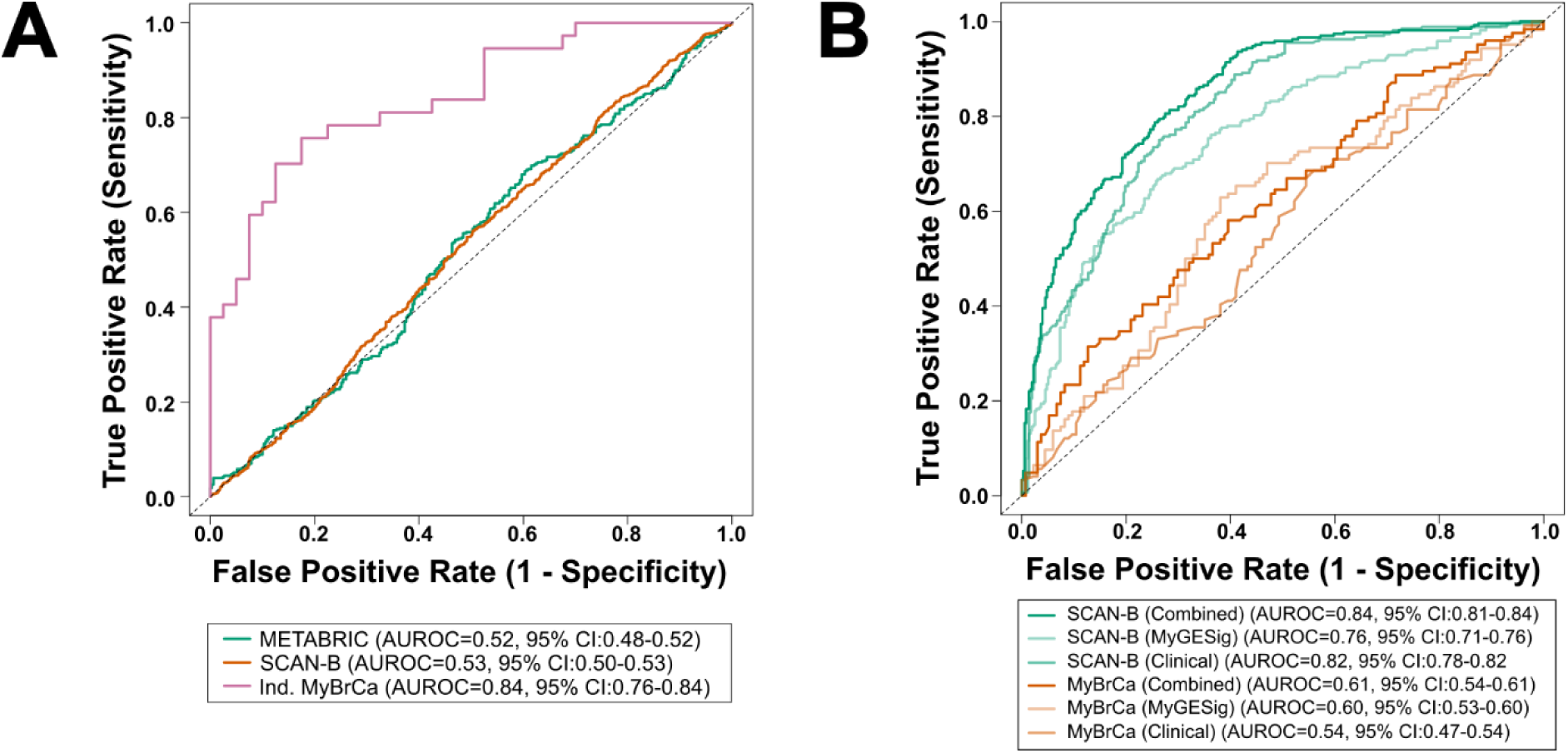
Discriminative performance of MyGESig gene signature across independent datasets. **A** ROC curves summarizing model performance in external validation datasets, including METABRIC, SCAN-B, and an independent MyBrCa cohort. The model was trained solely on gene expression from the MyBrCa dataset. **B** ROC curves comparing the performance of different SCANB70 models when evaluated on both its hold-out testing set (30% of the SCAN-B dataset) and the MyBrCa dataset. Models were trained using gene signatures, clinical information, or a combination of both.

To further assess the generalizability and reliability of the MyBrCa70 models, the SCAN-B and METABRIC cohorts were used as external validation cohorts. Models used in each validation were different as the MyBrCa70 models had to be retrained in the MyBrCa cohort, since the validation datasets varied in how certain clinical data were recorded, and only a subset of genes from the identified gene signature were present in these datasets. Specifically, the number of positive lymph nodes was recorded as a range in SCAN-B and was converted to a numeric value by taking the lower bound of the reported range. The microarray gene expression data from METABRIC only included 57 genes from the 77-gene signature, whereas the RNA-Seq dataset from SCAN-B included 59 genes. After retraining, the corresponding models performed poorly across all feature sets, with the median AUROC of 0.65, 0.52, and 0.66 (95% CIs: 0.60–0.64, 0.48–0.52, and 0.62–0.66) in METABRIC and 0.67, 0.53, and 0.62 (95% CIs: 0.64–0.67, 0.50–0.53, and 0.61–0.63) in SCAN-B, for when using clinical data, gene signatures, or both, respectively (Figure 6A, Supplementary Table 3).

Given the possible dataset-specific biases between the training and validation cohorts that can prevent machine learning model from capturing robust features that generalize beyond the training cohort, the respective external cohorts were used for model training (under the same 70:30 split; hereafter referred to as the SCANB70 and METABRIC70 models). The model’s discriminative performance improved for SCANB70, attaining the median AUROC of 0.85, 0.76, and 0.82 (95% CIs: 0.81–0.84, 0.72–0.76, and 0.78–0.82) when trained using clinical data, gene signatures, or both, respectively (Figure 6B, Supplementary Table 4). While a good discriminative performance for the clinical data suggests their contribution to the high AUROC in the combined model (SCANB70-Combined), the AUROC of 0.76 achieved by the SCANB70-MyGESig model in the hold-out SCAN-B testing dataset suggests that MyGESig gene set can moderately discriminate between prognosis groups when trained with cohort-specific data. In contrast, the same SCANB70 models performed poorly in the MyBrCa cohort, with the AUROC of 0.61, 0.60, and 0.54 (95% CIs: 0.54–0.61, 0.53–0.60, and 0.47–0.54), highlighting the poor generalizability of models trained with individual cohort data across cohorts. While moderate AUROC was still achieved in the METABRIC70-Combined model (0.71, 95% CI: 0.65-0.71), the METABRIC70-MyGESig demonstrated a poor discriminative performance when trained and tested in METABRIC (0.63, 95% CI: 0.58-0.63) (Supplementary Figure 5A). The weaker performance in this cohort is likely attributed to the difference in the platform used to measure gene expression (microarray vs RNA-seq). Taken together, these findings suggest that while the MyBrCa-trained model shows limited generalizability, MyGESig retains moderate discriminative ability, as demonstrated in the large SCAN-B dataset.

### 4.5 Assessment of the prognostic value of the MyGESig signature

Kaplan-Meier survival analysis comparing the IndMyBrCa30 models with PREDICT v3.0 revealed that both predictors were strongly associated with overall survival (p<0.0001), with the machine learning model demonstrating better separation of prognostic groups (Figure 7A). The hazard ratios (HRs) observed from Cox proportional hazard analysis for the IndMyBrCa30 models, PREDICT v3.0, as well as established clinical factors known for influencing overall survival, were summarized in a forest plot in Figure 7B. The results revealed that classification from both the IndMyBrCa30 models and PREDICT v3.0 was significantly associated with increased mortality, with the IndMyBrCa30-Combined model showing the strongest association (HR = 27.43, p-value < 0.01). Grade and HER2 positivity were not significantly associated with patient outcomes, whereas stage and HR positivity were significant. To demonstrate that the MyGESig signature has a prognostic value independent of clinical status, the IndMyBrCa30-MyGESig model was fit in a multivariable Cox model, controlling for stage and HR positivity. MyGESig remained significantly associated with patient outcomes (HR = 23.06, p-value < 0.001) (Table 4).

**Figure 7.**
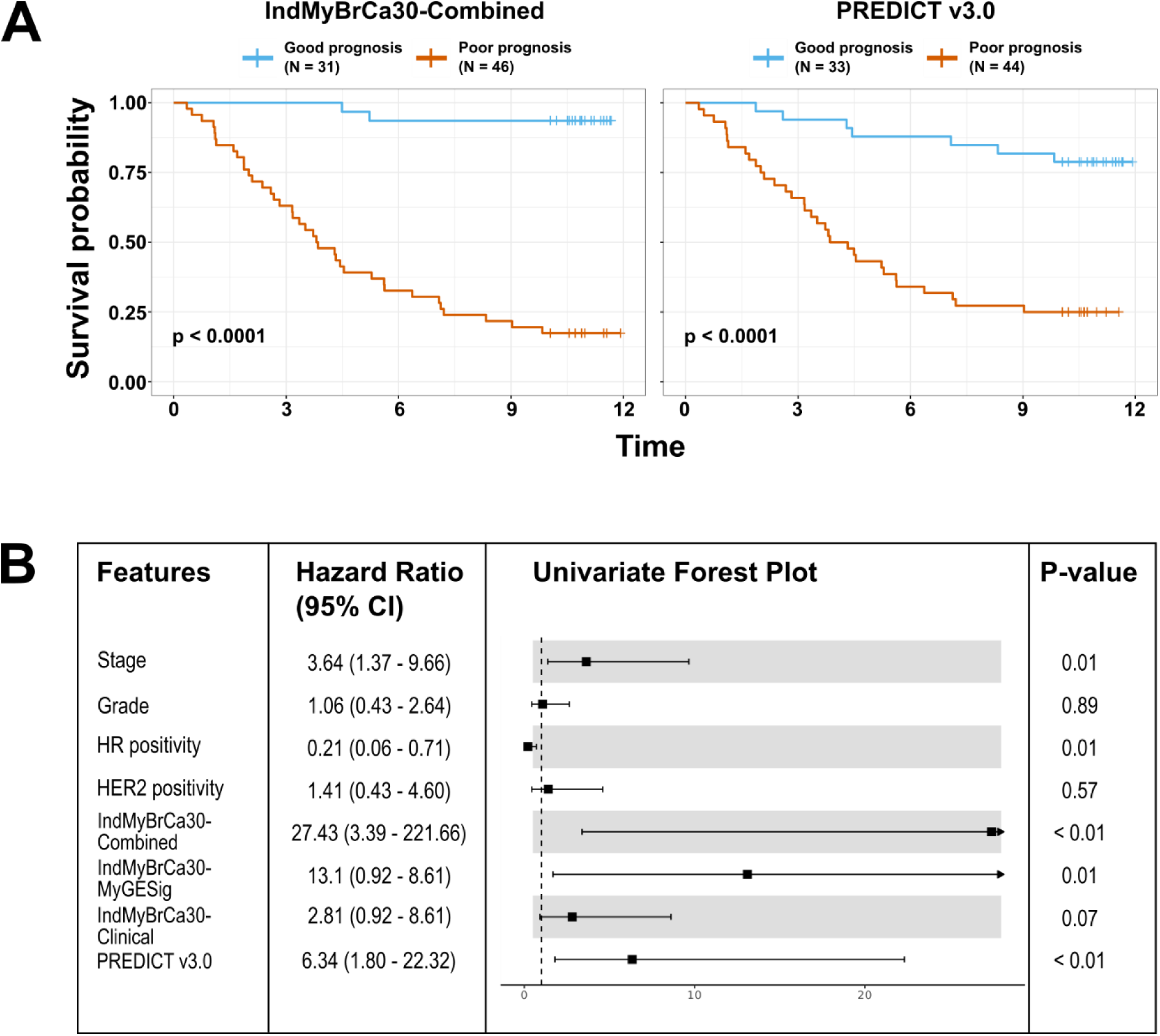
Evaluation of prognostic value of MyGESig compared to PREDICT v3.0 for the MyBrCa cohort. **A** Kaplan-Meier survival curves stratified by predicted prognosis; left, predictions generated by the prognostic model; right, predictions generated by PREDICT v3.0. **B** Forest plot summarizing hazard ratios (HRs) and 95% confidence intervals (CIs) derived from univariate Cox proportional hazards models for PREDICT, the IndMyBrCa30 models, and established clinical factors. Models trained on MyGESig, clinical information, or both were evaluated in the Cox framework.

**Table 4.** Multivariable Cox proportional hazards model results. Hazard ratios (HRs) with 95% confidence intervals (CIs) and p-values are shown for tumor stage, hormone receptor (HR) status, and classes assigned by the IndMyBrCa30-MyGESig model.

| Variables | Hazard ratio (95% CI) | P-value |
| --- | --- | --- |
| Stage | 2.44 (1.47-4.07) | <0.001 |
| HR status | 1.29 (0.64-2.61) | 0.474 |
| IndMyBrCa30-MyGESig | 23.06 (6.63-80.21) | <0.001 |

## 5 Discussion

In this study, the latest version of PREDICT v3.0 was validated in the MyBrCa cohort, and a prognostic gene signature was developed using transcriptomic profiles from the multi-ethnic Southeast Asian population. This gene signature, termed MyGESig, outperformed other established Western-derived gene signatures when used as input features in a machine learning-based model. Despite its poor cross-cohort performance, MyGESig demonstrated moderate-to-good discriminative performance when the models were trained and tested in the same dataset. Additionally, Cox proportional hazard analysis further confirmed the prognostic utility of this gene signature, with MyGESig remaining significantly associated with overall survival, independent of other established prognostic factors. Hence, given the moderate performance of PREDICT v3.0 in the MyBrCa cohort, this study highlights the prognostic value of a population-specific gene signature for improving prognostication in Malaysian breast cancer patients.

Compared to previous studies validating PREDICT v3.0 in Malaysian cohorts (Nik Ab Kadir et al., 2023; Wong et al., 2015), this study validated the most recently released version of PREDICT v3.0 using the largest sample size reported to date. Overall, PREDICT v3.0 slightly overestimated survival probabilities, with discrepancy from observed values increased in patients with lower overall survival and at longer time points (10- and 15-year predictions). The largest discrepancies in the observed versus estimated survival were found among patients with late-stage disease (Stage 3) and younger breast cancer patients—findings that were also corroborated by one of the earlier Malaysian-based studies (Wong et al., 2015). The PREDICT v3.0 tool has been previously validated in other populations, with varying calibration and discrimination performance (Magário et al., 2022; Nair et al., 2023; Stabellini et al., 2023; Wang et al., 2023; Zaguirre et al., 2021). These discrepancies have been attributed to differences in patient characteristics and clinical practices between the validation populations and the original population in which PREDICT v3.0 was developed.

The poor performance during cross-cohort validation of the models trained on MyBrCa does not necessarily indicate poor generalizability of the MyGESig gene set, as the gene set demonstrated moderate discriminative performance when trained and tested within the large SCAN-B dataset. Instead, it is the trained prognostic models (e.g., MyBrCa70-Combined and MyBrCa70-MyGESig), which incorporates the probabilistic RF and SVM algorithms, that may be susceptible to transferability issues. One potential reason for this lack of generalizability is the difference in patient demographics and characteristics between MyBrCa and the external datasets, leading to variation in both clinical features and gene expression that prevents the model from capturing a shared intrinsic pattern across both datasets (Lasko et al., 2024). A few solutions have been proposed to address this problem, including developing the model based on pooled cohort data. However, this will compromise the model’s performance in MyBrCa as a trade-off for generalizability. A more sophisticated approach is to separate the model into two components: a process model that identifies cohort-specific patterns to infer latent variables, and a disease model that uses these variables to make predictions (Lasko et al., 2024). While promising, this approach is beyond the scope of the current study and could be explored in future work. As with the findings of this study, it can be argued that good performance in external validation may not indicate universal applicability (Van Calster et al., 2023). Thus, this prognostic model may still have utility in the Malaysian population, especially considering the moderate performance of PREDICT. Hence, future work should focus on validating this model in the Malaysian population, particularly from other geographical regions.

This study has several limitations. First, the MyBrCa cohort is collected from participating private hospitals located in the urban region of Malaysia and includes a relatively high proportion of women of Chinese ethnicity compared to other ethnicities. Hence, this cohort may not fully represent the overall Malaysian population, particularly in the rural areas. Second, the assessment of PREDICT’s calibration in MyBrCa is limited by the high proportion of patients in the dataset with high overall survival; therefore, calibration cannot be reliably assessed at lower quintiles, particularly among patients with below 50% overall survival.

Additionally, KM’s estimation of observed survival may be affected by the high proportion of censoring in this cohort. Third, the gene signatures used for comparison —MammaPrint, Oncotype DX, and PAM50 as well as Eastern-based signatures such as GenesWell BCT and the signature from Cheng et al. (2016)—do not fully represent the actual commercial molecular assays (MammaPrint, Oncotype DX, Prosigna and GenesWell BCT). These signatures likely perform better when used within their prognostic models, which require data generated from their respective assays. Fourth, the independent MyBrCa dataset used for machine learning validation and Cox analysis consists of 77 patients who were excluded from the 70:30 split used for training and testing datasets. Hence, a larger sample size is needed to produce more robust estimates of AUROC and HR. Fifth, the machine learning models developed in this study may be subject to probability miscalibration, which could affect the accuracy of the predicted risk estimates and limit their clinical interpretability. As a potential future direction, post hoc calibration techniques such as Platt scaling or isotonic regression could be applied to improve the reliability of these probability estimates.

Taken together, the study presents a prognostic model that achieved 0.92 AUROC in predicting 10-year survival status in Malaysian breast cancer patients. The MyGESig signature has demonstrated both discriminative and prognostic value while complementing clinical information well in a machine learning setup. However, further development and validation within the Malaysian population are necessary to determine whether this model can perform reliably in real-world clinical settings.

## Data Availability

The whole exome sequencing and RNA-seq data included in this study are available in the European Genome-phenome Archive under Accession nos. EGAS00001006518 and EGAS00001004518. Access to controlled patient data will require the approval of the Data Access Committee. Further information is available from the corresponding author upon request.

## 6 Author Contributions

M.H.F.K. and J.-W.P. co-led the data analysis. M.H.F.K. wrote the manuscript. J.-W.P., S.C.C., S.-H.T., B.H.A., M.C.T., and W.K.H. drafted and reviewed the manuscript. J.-W.P., S.C.C., S.-H.T., B.H.A., M.C.T., and W.K.H. contributed to data analysis and interpretation. N.F.P. and J.P.-T. contributed to data analysis and drafted the manuscript. P.H., S.M.H., N.A.M.T., and C.-H.Y. contributed to sample collection and processing and data collection. Z.L.W. processed and cleaned the clinical data. The study was directed and co-supervised by S.C.C and J.-W.P. All authors read and approved the final version of the paper.

## 7 Acknowledgements

Cancer Research Malaysia receives charitable funding from the Scientex Foundation, Estée Lauder Companies, Vistage Malaysia, Yayasan PETRONAS, and Yayasan Sime Darby which contributed to the funding of this study. Funding was also provided by a research grant from the Newton-Ungku Omar Fund (MRC Ref: MR/P012442/1) to SFC and SHT. OMR, CC, and SFC also receive funding from Cancer Research UK. All genomics work was undertaken by the Genomics Core Facility CRUK Cambridge Institute.

## 9 Ethics declaration

Patient recruitment and sample collection was reviewed and approved by the Independent Ethics Committee, Ramsay Sime Darby Health Care (Reference nos: 201109.4 and 201208.1), as well as the Medical Ethics Committee of the University Malaya Medical Centre (Reference no: 842.9). Written informed consent to participation in research was given by each individual patient.

## Supplementary Figures

**Supplementary Figure 1.**
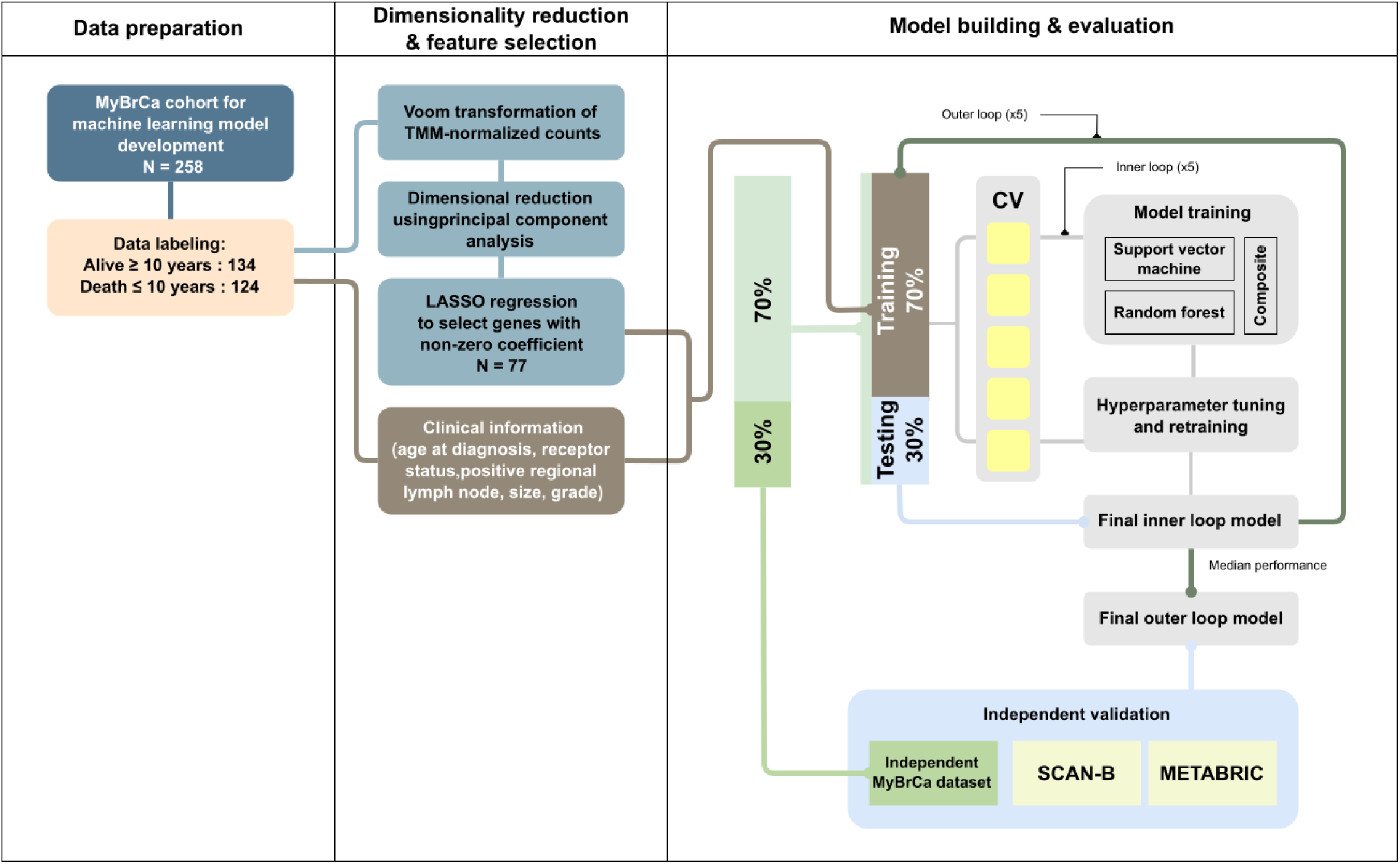
Diagram summarizing the development of the prognostic model. For validation on the independent MyBrCa cohort, 30% of the cohort was held out as an independent dataset, while the remaining 70% was used for model training and testing, following the same 7:3 stratified split. Note that only the model used for this independent validation was trained on the reduced second-split dataset; all other models were trained using the original single-split dataset.

**Supplementary Figure 2.**
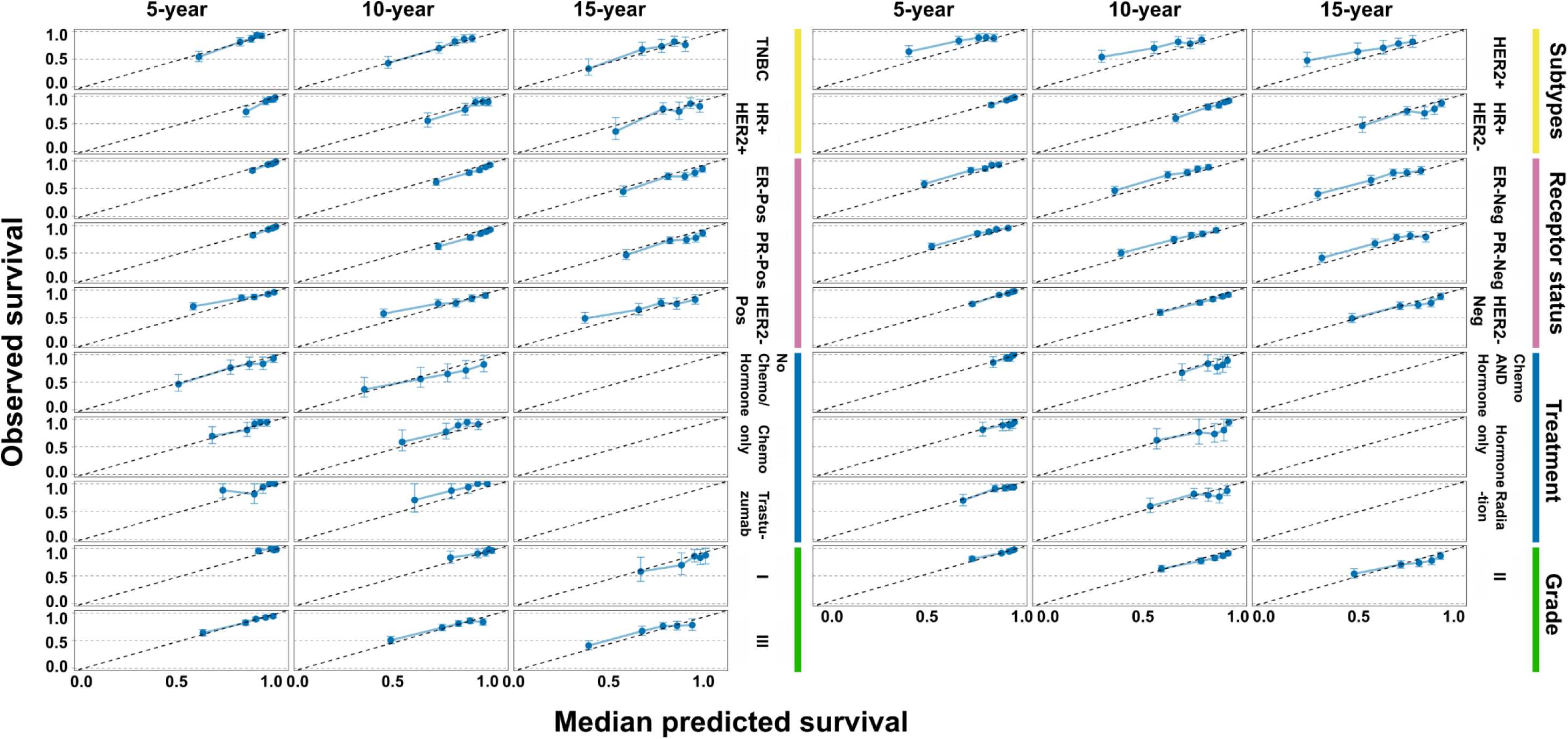
Evaluation of PREDICT’s performance across different subgroups of patients in the MyBrCa cohort. Calibration plots comparing the predicted versus observed survival in various subgroups of patients in the MyBrCa cohort. Predictions were binned into quintiles for visualization. The diagonal line represents perfect calibration performance. Only subgroups of known clinical variables were selected for visualization. Calibration at the 15-year timepoint could not be computed for the treatment subgroups, due to the absence of patients with 15-year follow-up.

**Supplementary Figure 3.**
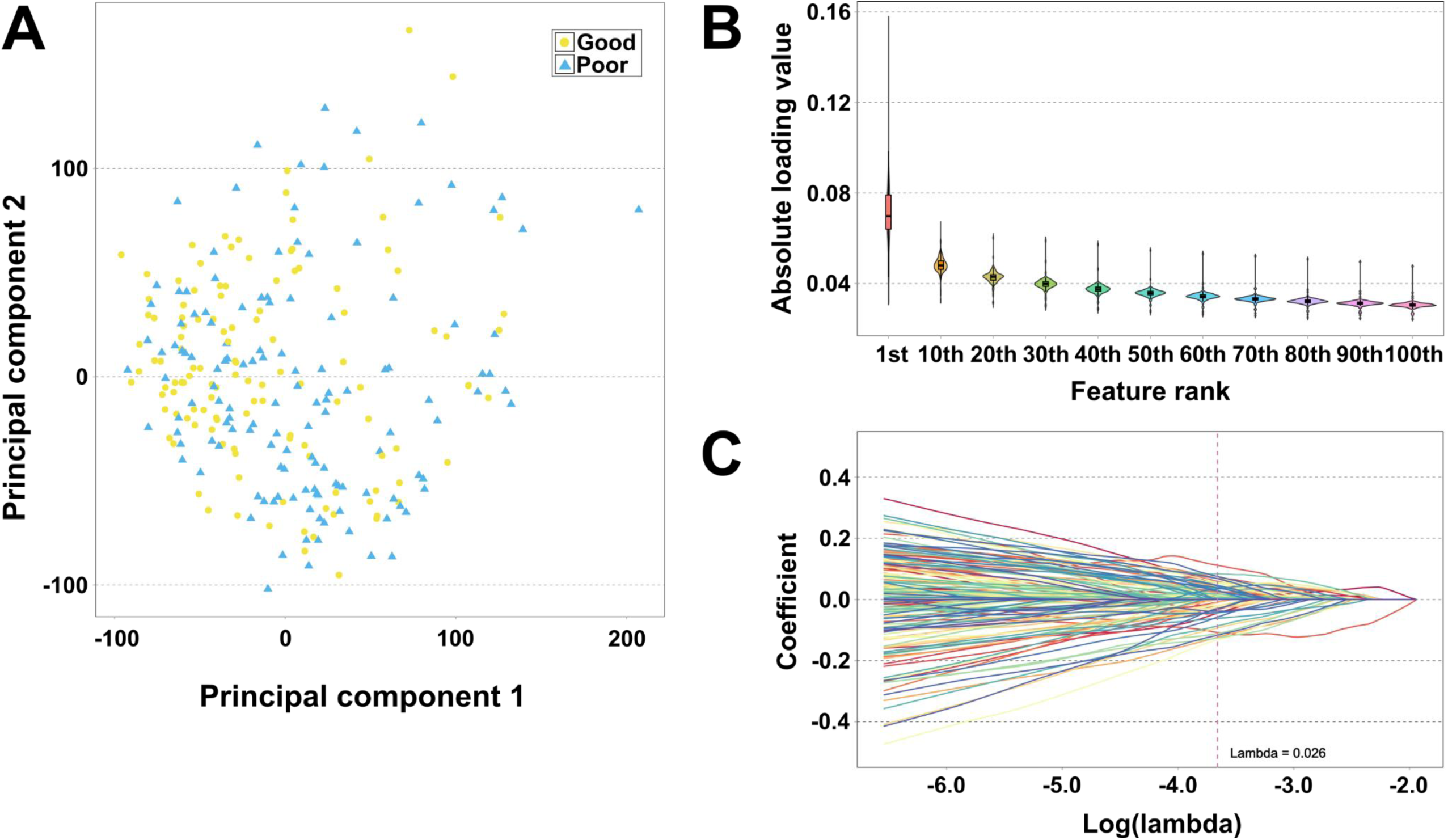
Dimensionality reduction in the transcriptome dataset for subsequent feature selection. **A** Principal component analysis (PCA) plot of the transcriptomic dataset for 258 MyBrCa patients, coloured by prognosis groups. No clear clustering by prognosis was observed. The top 30 genes with the highest absolute loadings in each principal component were retained for further analysis. **B** Distribution of absolute loading values across all principal components at different cut-off points, with genes ranked in descending order of absolute loading. **C** Plot of LASSO regression coefficients against log-transformed lambda values, illustrating the shrinkage of coefficients to zero as lambda increases. The final lambda value selected was 0.026.

**Supplementary Figure 4.**
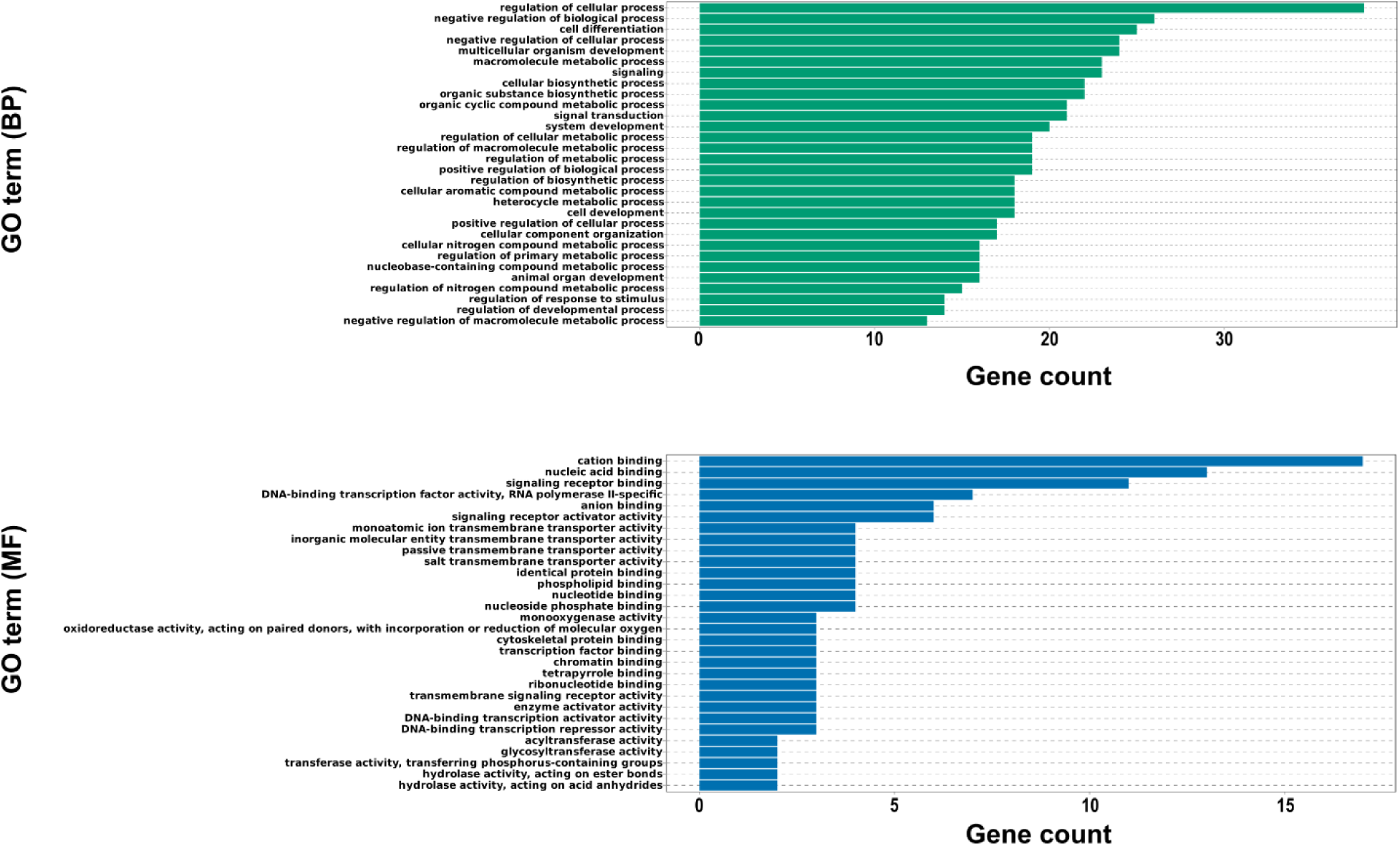
Gene Ontology (GO) annotations for genes in the MyGESig signature. Bar plots showing the top 30 most frequently assigned GO terms in the MyGESig gene signature. No term was significantly enriched in the gene set.

**Supplementary Figure 5.**
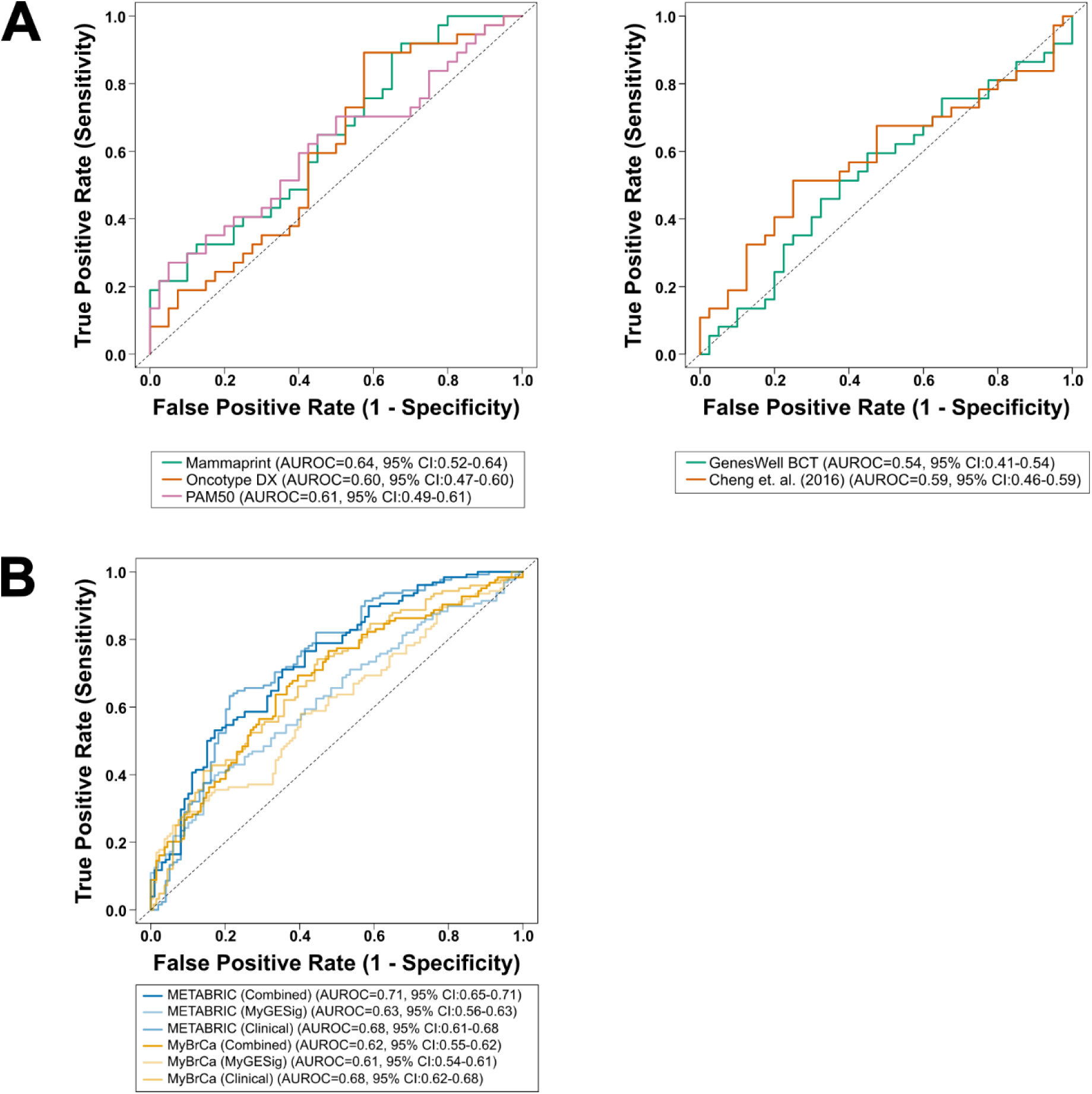
Comparison of model performance across different gene signatures and training datasets. **A** ROC curves comparing the performance of models when established Western-based(Mammaprint, Oncotype DX, and PAM50) (left) and Eastern-based gene signatures (GenesWell BCT and signature from Cheng et. al. (2016)) were used as features. **B** ROC curves comparing the performance of different METABRIC70 models when evaluated on both its hold-out testing set (30% of the METABRIC dataset) and the MyBrCa dataset. Models were trained using gene signatures, clinical information, or a combination of both.

